# Methaemoglobin as a surrogate marker of primaquine antihypnozoite activity in *Plasmodium vivax* malaria: a systematic review and individual patient data meta-analysis

**DOI:** 10.1101/2024.05.08.24307041

**Authors:** Ihsan Fadilah, Robert J Commons, Nguyen Hoang Chau, Cindy S Chu, Nicholas PJ Day, Gavin CKW Koh, Justin A Green, Marcus VG Lacerda, Alejandro Llanos-Cuentas, Erni J Nelwan, Francois Nosten, Ayodhia Pitaloka Pasaribu, Inge Sutanto, Walter RJ Taylor, Kamala Thriemer, Ric N Price, Nicholas J White, J Kevin Baird, James A Watson

## Abstract

**Background:** The 8-aminoquinolines, primaquine and tafenoquine, are the only available drugs for the radical cure of *Plasmodium vivax* hypnozoites. Prior evidence suggests that there is dose-dependent 8-aminoquinoline induced methaemoglobinaemia and that higher methaemoglobin concentrations are associated with a lower risk of *P. vivax* recurrence. We undertook a systematic review and individual patient data meta-analysis to examine the utility of methaemoglobin as a surrogate endpoint for 8-aminoquinoline antihypnozoite activity to prevent *P. vivax* recurrence.

**Methods:** We conducted a systematic search of Medline, Embase, Web of Science, and the Cochrane Library, from 1 January 2000 to 29 September 2022 inclusive, of prospective clinical efficacy studies of acute, uncomplicated *P. vivax* malaria mono-infections treated with radical curative doses of primaquine. The day 7 methaemoglobin concentration was the primary surrogate outcome of interest. The primary clinical outcome was the time to first *P. vivax* recurrence between day 7 and day 120 after enrolment. We used multivariable Cox proportional-hazards regression with site random-effects to characterise the time to first recurrence as a function of the day 7 methaemoglobin percentage (log_2_ transformed), adjusted for the partner schizontocidal drug, the primaquine regimen duration as a proxy for the total primaquine dose (mg/kg), the daily primaquine dose (mg/kg), and other factors. The systematic review protocol was registered with PROSPERO (CRD42023345956).

**Findings:** We identified 219 *P. vivax* efficacy studies, of which eight provided relevant individual-level data from patients treated with primaquine; all were randomised, parallel arm clinical trials assessed as having low or moderate risk of bias. In the primary analysis dataset, there were 1747 G6PD-normal patients enrolled from 24 study sites across 8 different countries (Indonesia, Brazil, Vietnam, Thailand, Peru, Colombia, Ethiopia, India). We observed an increasing dose-response relationship between the daily weight-adjusted primaquine dose and day 7 methaemoglobin level. For a given primaquine dose regimen, an observed doubling in day 7 methaemoglobin percentage was associated with an estimated 30% reduction in the risk of vivax recurrence (adjusted hazard ratio = 0.70; 95% CI = [0.57, 0.86]; p = 0.0005). These pooled estimates were largely consistent across the study sites. Using day 7 methaemoglobin as a surrogate endpoint for recurrence would reduce required sample sizes by approximately 40%.

**Conclusions:** For a given primaquine regimen, higher methaemoglobin on day 7 was associated with a reduced risk of *P. vivax* recurrence. Under our proposed causal model, this justifies the use of methaemoglobin as a surrogate endpoint for primaquine antihypnozoite activity in G6PD normal patients with *P. vivax* malaria.

## Introduction

The human malaria parasites *Plasmodium vivax* and *Plasmodium ovale* are characterised by their ability to form dormant liver stage parasites called hypnozoites, which activate weeks to months later to cause relapsing bloodstream infection [1]. *P. vivax* is the most geographically widespread cause of human malaria and is a major challenge in malaria elimination. Relapse contributes substantially to the overall burden of symptomatic vivax malaria, causing over 75% of all symptomatic infections [2]. Preventing relapse is crucial for eliminating the burden of vivax malaria morbidity and mortality.

The only drugs available for radical cure (killing latent hypnozoites) are primaquine and tafenoquine. Both are thought to be prodrugs [3, 4], necessitating metabolic activation to produce hypnozonticidal activity. The precise mechanism and active metabolites of primaquine and tafenoquine remain unknown [5]. Patients with vivax malaria, who receive an equivalent 8-aminoquinoline dose per body weight, may variably metabolise the drug leading to varying risks of later relapse. Some of this variation in biotransformation is due to polymorphisms in cytochrome *P450 (CYP) 2D6* [3] and poorer metabolisers are associated with higher relapse rates [6, 7].

Both primaquine and tafenoquine cause predictable increases in blood methaemoglobin resulting from the action of their oxidative metabolites [8]. This involves a reversible increase in the conversion rate of intra-erythrocytic reduced (Fe^++^) haem iron in haemoglobin to its oxidised (Fe^+++^) form [8]. Following daily administration of primaquine or single dose administration of tafenoquine, blood methaemoglobin gradually increases, reaching a peak concentration after approximately one week [9]. It has been hypothesised that the same oxidative metabolites responsible for methaemoglobinaemia are also responsible for inducing haemolysis in glucose-6-phosphate dehydrogenase (G6PD) deficiency, killing mature gametocytes of *P. falciparum*, and killing liver stage hypnozoites [8]. Early experiments with primaquine and its analogues indicated that there were greater increases in blood methaemoglobin for molecules which had improved radical-cure efficacy [8]. Two recent pharmacometric studies estimated an adjusted proportional reduction in the risk of vivax recurrence of approximately 10% for primaquine [10] and 20% for tafenoquine [4] for each additional percentage-point increase in day 7 methaemoglobin. Based on these results, it is hypothesised that increases in methaemoglobin may serve as a proxy for 8-aminoquinoline antihypnozoite activity and, as such, a potential surrogate endpoint for clinical trials to quantify the antirelapse efficacy of 8-aminoquinoline drugs in vivax malaria. A surrogate endpoint is a patient characteristic, such as a biomarker, intended to substitute for a clinical outcome [11]– specifically vivax recurrence in this context. To validate surrogacy, high-quality studies need to demonstrate that a putative biomarker is affected by the drug intervention and that drug-induced change in the biomarker level can predict the effect on the outcome of interest [12, 13].

We aimed to examine the utility of methaemoglobin as a surrogate endpoint for vivax recurrence using all available data in a pooled individual patient data meta-analysis involving *P. vivax* patients from multiple countries treated with primaquine.

## Methods

### Search strategy and selection criteria

We conducted a systematic search of Medline, Embase, Web of Science, and the Cochrane Library based on an existing living systematic review [14] of prospective clinical efficacy studies of acute, uncomplicated *P. vivax* malaria mono-infections treated with primaquine. We included randomised therapeutic trials and prospective cohort studies published between 1 January 2000 and 29 September 2022 inclusive, in any language, with a minimum active follow-up of 42 days that recorded methaemoglobin data (at baseline and at least once in the first week of follow-up between day 5 and day 9) following daily primaquine administration given over multiple days. Only G6PD normal individuals (i.e., ≥30% G6PD activity or a negative qualitative test) were included in the analysis. In G6PD deficient individuals, primaquine administration does not lead to the same methaemoglobin increases [15].

Studies were included if the primaquine regimen was administered within the first three days of schizontocidal treatment. Search terms are provided in Supporting Information (List S1). This systematic review was conducted by two reviewers (IF and RJC), with discrepancies resolved through discussion. The review protocol was registered with PROSPERO (CRD42023345956).

### Data pooling

The corresponding authors and/or principal investigators of eligible studies that met the study criteria were invited through direct email to contribute their individual patient data. Relevant data from unpublished studies were requested wherever possible. Shared data were uploaded to the Worldwide Antimalarial Resistance Network (WWARN) repository for curation and standardisation, utilising the IDDO SDTM Implementation Guide [16]. We excluded individual patients with missing information on age, sex, body weight, baseline parasite density, primaquine regimen, or schizontocidal treatment. Patients with severe malaria, pregnancy, mixed-species infection, or those who received adjunctive antimalarials after the initial schizontocidal treatment were also excluded.

All studies included in our meta-analysis provided pseudonymised individual data and had obtained ethical approvals from the corresponding site of origin. Therefore, additional ethical approval was not required for the current analysis, as per the Oxford Tropical Research Ethics Committee. We adhered to the PRISMA-IPD guidelines [17] for reporting this systematic review and individual patient data meta-analysis (checklist available in Supporting Information, Table S1).

### Outcomes

The primary clinical outcome was the time to first *P. vivax* recurrence (i.e., any episode of *P. vivax* parasitaemia irrespective of symptoms) between day 7 (the starting point for prediction and follow-up) and day 120 [4] after the initial primaquine administration.

The secondary outcomes were the binary outcome of any *P. vivax* recurrence between day 7 and day 120 after primaquine initiation and the maximum absolute change in haemoglobin concentration from day 0 to days 2–3 (expected days of the lowest haemoglobin level [18]) following the start of any antimalarial treatment. In addition to being the main predictor of interest in the statistical model, we also specified the day 7 methaemoglobin concentration as an outcome and modelled this biomarker as a function of daily mg (base) per kilogram primaquine dose.

### Data analysis

We presented study-level summary statistics to highlight sample characteristics and potential heterogeneity across the included studies. The daily distributions of methaemoglobin levels, stratified by schizontocidal drug and primaquine regimen (low total dose 14-day, high total dose 7-day), were plotted to illustrate the temporal dynamics of primaquine-induced methaemoglobin production during radical cure treatment.

The primary predictor of interest (surrogate outcome) was the day 7 methaemoglobin concentration, expressed as a percentage of the total haemoglobin concentration. Day 7 was prespecified and typically, methaemoglobin concentrations peak after approximately a week of commencing the daily primaquine regimens. All studies measured methaemoglobin by transcutaneous pulse CO-oximetry. The day 7 methaemoglobin percentage (log_2_ transformed) was included in the statistical model as a continuous variable. If the day 7 methaemoglobin percentage was recorded as zero, this remained untransformed (which assumed a methaemoglobin of 1%–approximately the physiological level). Zero recordings are likely to represent mis-readings of the analytical machine.

We proposed a causal directed acyclic graph for this analysis to guide model specification and aid interpretation of results (Fig 1). Missing day 7 methaemoglobin percentages were linearly imputed using levels measured within ±2 days. If only one measurement was available, then the imputation assumed a constant (i.e., the single value observed was used). If no measurements were available within this timeframe (day 5 to day 9), the patient was excluded from the analysis.

**Figure 1.**
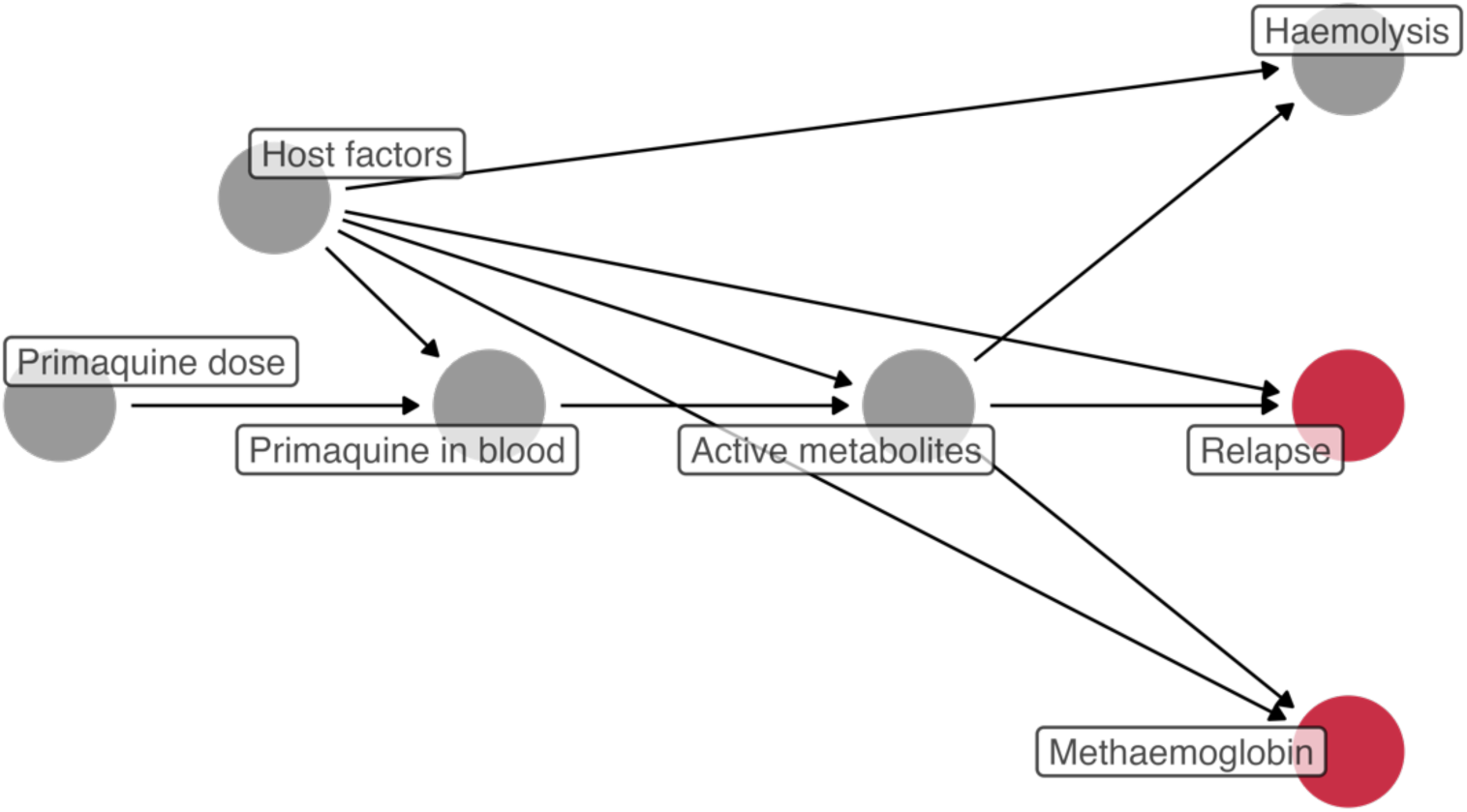
Directed acyclic graph showing our hypothesised causal relationships between primaquine-induced changes in blood methaemoglobin and *P. vivax* relapse. Red nodes represent the outcomes of interest: relapse and blood methaemoglobin (measured on day 7, for example), between which the association was estimated. Under this causal model, blood methaemoglobin is a proxy measurement for the hypnozontocidal activity of primaquine (but not on the causal pathway mediating the effect of primaquine on relapse). Host factors include but not limited to patient’s genetics (e.g., those related to *CYP2D6* and *G6PD*), behaviours, age, immunity to *P. vivax*, and geographical location.

In the main analysis, patients were right censored at the time of the first recurrent vivax parasitaemia (outcome), any malaria parasitaemia, loss to follow up, blood smear gap of >60 days, or the last day of study; whichever occurred first. We used multivariable, random-effects Cox proportional-hazards regression to model the time to first recurrence as a function of the day 7 methaemoglobin percentage (log_2_ transformed) under a one-stage individual patient data meta-analysis framework. This model adjusted for daily mg/kg primaquine dose, primaquine duration (a proxy for total mg/kg primaquine dose), within-site and across-site linear interactions [19] between daily mg/kg primaquine dose and primaquine duration, age, sex, schizontocidal drug, and baseline parasite density (natural-log transformed). A random intercept and a random slope for day 7 methaemoglobin concentration were included to account for between-site effect-heterogeneity. Linearity and proportional-hazards assumptions were checked. The adjusted hazard ratio can be interpreted as the estimated predictive effect of each doubling in day 7 methaemoglobin percentage, over and above the adjustment factors.

To compare our estimates with those from previous studies [4, 10], we also specified the day 7 methaemoglobin concentration on its original scale and refitted the main survival analysis model. Additionally, we separately (by study and primaquine regimen) fit a more parsimonious Cox proportional-hazards model that adjusted for daily mg/kg primaquine dose and pooled the estimates obtained from all the resulting clusters using a two-stage individual-patient data meta-analysis approach. A similar model specification to the one-stage approach that included a few more adjustment factors was not possible as the data were sparse (i.e., few recurrences). A forest plot was constructed to visualise the results under the common-effect and random-effects models.

We estimated the adjusted predictive effect of the day 7 methaemoglobin percentage (log_2_ transformed) on the odds of vivax recurrence using multivariable, random-effects binary logistic regression. This model was limited to patients with at least 120 days of follow-up and adjusted for daily mg/kg primaquine dose, primaquine duration, within-site and across-site linear interactions between daily mg/kg primaquine dose and primaquine duration. A random intercept for study site was specified. The association between the maximum absolute change in haemoglobin concentration from day 0 to days 2–3 and the day 7 methaemoglobin percentage was estimated using multivariable, random-effects linear regression. This model included baseline haemoglobin concentration, daily mg/kg primaquine dose, age, sex, schizontocidal drug, baseline parasite density (natural-log transformed) as common-effect covariates, and a random intercept and slope for study site and daily mg/kg primaquine dose, respectively. This model was restricted to patients who started primaquine treatment on day 0. If a haemoglobin measurement was missing, haematocrit was used to impute the haemoglobin concentration using the formula haemoglobin = (haematocrit − 5.62) ÷ 2.60, where haematocrit was measured in percent and haemoglobin was measured in grams per decilitre [20]. If haematocrit remained missing, these patients were excluded. We also estimated the association of daily mg/kg primaquine dose and day 7 methaemoglobin percentage using a random-effects linear model, allowing for a random intercept and slope for study site and daily mg/kg primaquine dose, respectively.

We provide illustrative sample-size calculations to demonstrate how our findings could contribute to making future studies of drug discovery or regimen optimisation in *P. vivax* more efficient by using blood methaemoglobin as a surrogate outcome. We estimated that a 0.5-mg/kg increase in daily primaquine dose results in a 0.39 increase in the log_2_ day 7 methaemoglobin (i.e. a 30% increase). We estimated the standard deviation of the log_2_ day 7 methaemoglobin level conditional on the daily mg/kg primaquine dose from the pooled data. Assuming a normal distribution for the log_2_ day 7 methaemoglobin conditional on the daily dose allows for a simple calculation of the required sample size (based on a test for a difference between two normal distributions).

Risk of bias related to individual studies was evaluated using the Quality in Prognosis Studies (QUIPS) tool [21] adapted to the current analysis (signalling questions are provided in Supporting Information, List S2). Statistical analysis followed a prespecified plan [22] and was conducted using R Statistical Software (version 4.3.0).

## Findings

### Study and patient selection

We identified 219 *P. vivax* efficacy studies published between 1 January 2000 and 29 September 2022 (Fig 2). After review, 206 studies were excluded, leaving 13 studies eligible for the pooled analysis. Eight of these studies (8/13, 62%) provided individual level data from patients treated with primaquine; all were randomised, parallel arm clinical trials and assessed as having low risk of bias (Table S2). Individual level data were available for 4122 patients from these eight trials, of whom 1747 (42%) satisfied the inclusion and exclusion criteria for our analysis (primary dataset). These patients were enrolled from 24 study sites across 8 different countries (Indonesia, Brazil, Vietnam, Thailand, Peru, Colombia, Ethiopia, India; Fig S1). Most patients had a follow-up of at least 120 days since commencing primaquine radical cure treatment (1344, 77%–secondary sub-dataset) and had haemoglobin concentration (or haematocrit) measured on day 0 of antimalarial treatment (1360, 78%–haemolysis sub-dataset).

**Figure 2.**
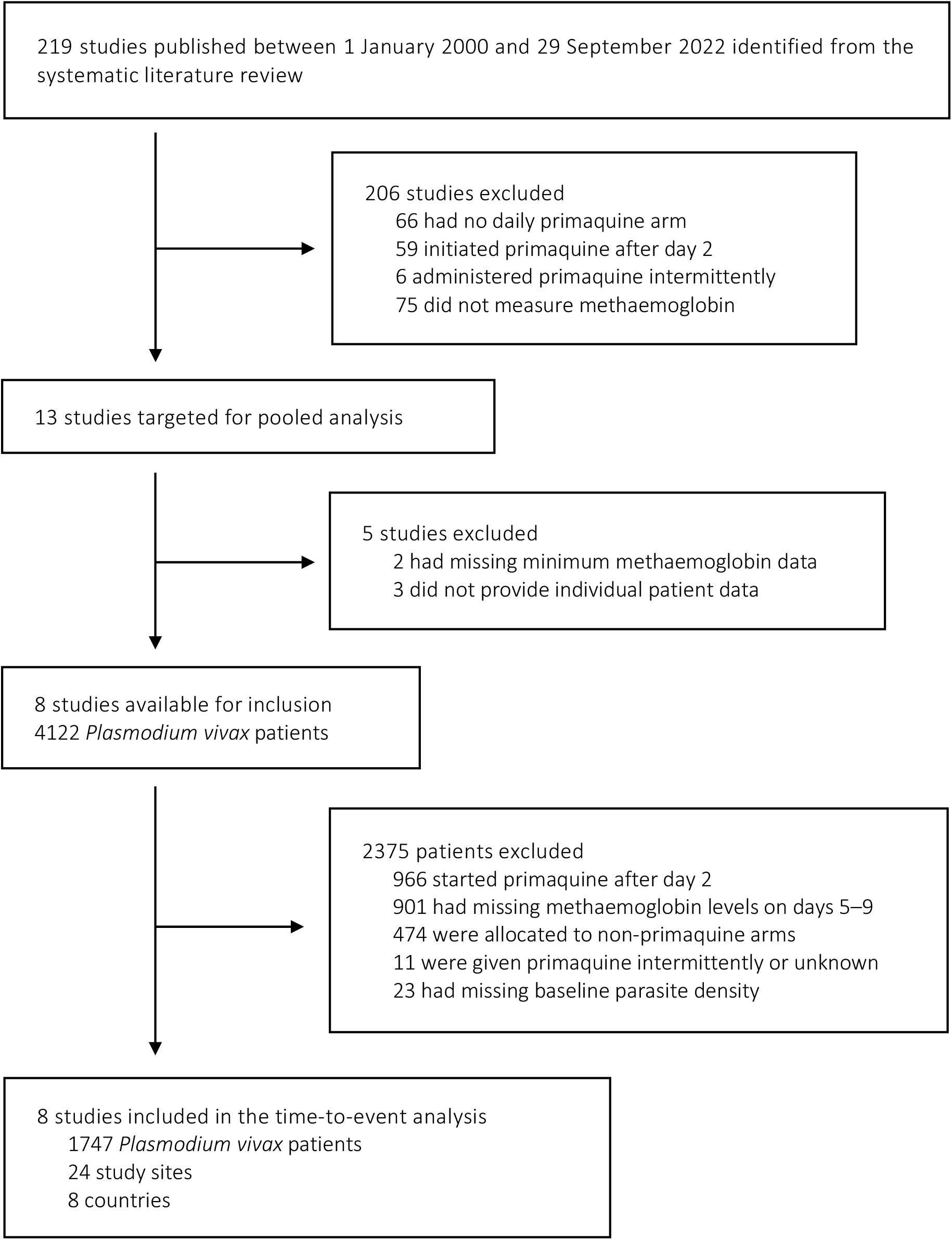
Study and patient selection. Databases systematically searched were from Medline, Embase, Web of Science, and the Cochrane Library. Patients included in the secondary analyses were subsets of the patients in the primary, time-to-event analysis.

### Patient characteristics

In the primary analysis (n = 1747 patients), the median age was 20 years (interquartile range [IQR]: 12 to 32), with 89 patients (5.1%) younger than 5 years. Overall, most patients were male (1116, 64%), resided in the Asia-Pacific region (1620, 93%), and were from locations with frequent relapse periodicity (i.e., a median interval from the first acute episode to relapse of less than 47 days [23]; 1614, 92%) and moderate transmission intensity (i.e., 1 to 9 cases per 1000 person-years [24]; 1325, 76%). The majority of patients (1138, 65%) were treated with artemisinin-combination therapies (ACT), started primaquine on day of enrolment (day 0; 1496, 86%), and took daily primaquine over 14 days (1194, 68%). The overall median daily-dose of primaquine was 0.52 mg/kg (IQR: 0.38 to 0.95, Fig S2 shows the weight-adjusted dose distribution by primaquine duration). Primaquine administration was fully supervised in most studies (i.e., all doses were directly observed; 1571, 90%). Further details on baseline patient characteristics are summarised in Table 1. Eligible studies that were not included [25–29] in the analysis tended to have a shorter duration of follow-up but were otherwise similar in other characteristics (Tables S3–5).

**Table 1.** Demographic and patient characteristics at baseline.

| Characteristic | Overall | Study |  |  |  |  |  |  |  |
| --- | --- | --- | --- | --- | --- | --- | --- | --- | --- |
|  |  | Pasaribu 2013 [42] | Sutanto 2013 [39] | Llanos-Cuentas 2014 [36] | Nelwan 2015 [40] | Chu 2019 [9] | Lacerda 2019 [35] | Llanos-Cuentas 2019 [37] | Taylor 2019 [43] |
| **Number of patients** | 1,747 | 303 | 38 | 50 | 120 | 578 | 42 | 84 | 532 |
| **Region** |  |  |  |  |  |  |  |  |  |
| Asia-Pacific | 1,620 (93%) | 303 (100%) | 38 (100%) | 22 (44%) | 120 (100%) | 578 (100%) | 4 (9.5%) | 23 (27%) | 532 (100%) |
| Americas | 124 (7.1%) | 0 (0%) | 0 (0%) | 28 (56%) | 0 (0%) | 0 (0%) | 35 (83%) | 61 (73%) | 0 (0%) |
| Africa | 3 (0.2%) | 0 (0%) | 0 (0%) | 0 (0%) | 0 (0%) | 0 (0%) | 3 (7.1%) | 0 (0%) | 0 (0%) |
| **Relapse periodicity<sup>§</sup>** |  |  |  |  |  |  |  |  |  |
| Low | 133 (7.6%) | 0 (0%) | 0 (0%) | 34 (68%) | 0 (0%) | 0 (0%) | 38 (90%) | 61 (73%) | 0 (0%) |
| High | 1,614 (92%) | 303 (100%) | 38 (100%) | 16 (32%) | 120 (100%) | 578 (100%) | 4 (9.5%) | 23 (27%) | 532 (100%) |
| **Transmission intensity<sup>#</sup>** |  |  |  |  |  |  |  |  |  |
| Low | 262 (15%) | 0 (0%) | 0 (0%) | 16 (32%) | 120 (100%) | 0 (0%) | 4 (9.5%) | 17 (20%) | 105 (20%) |
| Moderate | 1,325 (76%) | 303 (100%) | 0 (0%) | 6 (12%) | 0 (0%) | 578 (100%) | 0 (0%) | 11 (13%) | 427 (80%) |
| High | 160 (9.2%) | 0 (0%) | 38 (100%) | 28 (56%) | 0 (0%) | 0 (0%) | 38 (90%) | 56 (67%) | 0 (0%) |
| **Age (years)** | 20 (12, 32) | 13 (9, 25) | 27 (25, 29) | 34 (26, 46) | 28 (25, 31) | 20 (13, 32) | 38 (24, 47) | 36 (25, 50) | 15 (10, 29) |
| **Age (years)** |  |  |  |  |  |  |  |  |  |
| <5 | 89 (5.1%) | 26 (8.6%) | 0 (0%) | 0 (0%) | 0 (0%) | 24 (4.2%) | 0 (0%) | 0 (0%) | 39 (7.3%) |
| ≥5 and <15 | 534 (31%) | 144 (48%) | 0 (0%) | 0 (0%) | 0 (0%) | 168 (29%) | 0 (0%) | 0 (0%) | 222 (42%) |
| ≥15 | 1,124 (64%) | 133 (44%) | 38 (100%) | 50 (100%) | 120 (100%) | 386 (67%) | 42 (100%) | 84 (100%) | 271 (51%) |
| **Male** | 1,116 (64%) | 169 (56%) | 38 (100%) | 35 (70%) | 120 (100%) | 363 (63%) | 30 (71%) | 52 (62%) | 309 (58%) |
| **Body weight (kg)** | 48 (30, 59) | 35 (21, 50) | 65 (59, 72) | 59 (49, 68) | 69 (63, 74) | 47 (33, 54) | 64 (55, 72) | 63 (55, 70) | 43 (24, 54) |
| **Schizontocidal drug** |  |  |  |  |  |  |  |  |  |
| Artesunate/Amodiaquine | 146 (8.4%) | 146 (48%) | 0 (0%) | 0 (0%) | 0 (0%) | 0 (0%) | 0 (0%) | 0 (0%) | 0 (0%) |
| Artesunate/Pyronaridine | 60 (3.4%) | 0 (0%) | 0 (0%) | 0 (0%) | 60 (50%) | 0 (0%) | 0 (0%) | 0 (0%) | 0 (0%) |
| Dihydroartemisinin/Piperaquine | 932 (53%) | 157 (52%) | 0 (0%) | 0 (0%) | 60 (50%) | 290 (50%) | 0 (0%) | 0 (0%) | 425 (80%) |
| Quinine | 38 (2.2%) | 0 (0%) | 38 (100%) | 0 (0%) | 0 (0%) | 0 (0%) | 0 (0%) | 0 (0%) | 0 (0%) |
| Chloroquine | 571 (33%) | 0 (0%) | 0 (0%) | 50 (100%) | 0 (0%) | 288 (50%) | 42 (100%) | 84 (100%) | 107 (20%) |
| **Primaquine duration** |  |  |  |  |  |  |  |  |  |
| 7 days | 553 (32%) | 0 (0%) | 0 (0%) | 0 (0%) | 0 (0%) | 289 (50%) | 0 (0%) | 0 (0%) | 264 (50%) |
| 14 days | 1,194 (68%) | 303 (100%) | 38 (100%) | 50 (100%) | 120 (100%) | 289 (50%) | 42 (100%) | 84 (100%) | 268 (50%) |
| **Primaquine start** |  |  |  |  |  |  |  |  |  |
| Day 0 | 1,496 (86%) | 303 (100%) | 27 (71%) | 0 (0%) | 120 (100%) | 511 (88%) | 3 (7.1%) | 0 (0%) | 532 (100%) |
| Day 1 | 181 (10%) | 0 (0%) | 9 (24%) | 49 (98%) | 0 (0%) | 0 (0%) | 39 (93%) | 84 (100%) | 0 (0%) |
| Day 2 | 70 (4.0%) | 0 (0%) | 2 (5.3%) | 1 (2.0%) | 0 (0%) | 67 (12%) | 0 (0%) | 0 (0%) | 0 (0%) |
| **Primaquine daily dose (mg/kg)** | 0.52 (0.38, 0.95) | 0.31 (0.27, 0.34) | 0.51 (0.47, 0.59) | 0.25 (0.22, 0.30) | 0.53 (0.47, 0.60) | 0.87 (0.50, 1.00) | 0.23 (0.21, 0.27) | 0.25 (0.23, 0.29) | 0.66 (0.53, 1.03) |
| **Primaquine daily dose (mg/kg)** |  |  |  |  |  |  |  |  |  |
| <0.375 | 436 (25%) | 253 (83%) | 0 (0%) | 49 (98%) | 0 (0%) | 4 (0.7%) | 42 (100%) | 83 (99%) | 5 (0.9%) |
| ≥0.375 and <0.75 | 760 (44%) | 50 (17%) | 38 (100%) | 1 (2.0%) | 120 (100%) | 282 (49%) | 0 (0%) | 1 (1.2%) | 268 (50%) |
| ≥0.75 | 551 (32%) | 0 (0%) | 0 (0%) | 0 (0%) | 0 (0%) | 292 (51%) | 0 (0%) | 0 (0%) | 259 (49%) |
| **Primaquine dose calculation** |  |  |  |  |  |  |  |  |  |
| Actual dosing | 1,324 (76%) | 0 (0%) | 38 (100%) | 50 (100%) | 0 (0%) | 578 (100%) | 42 (100%) | 84 (100%) | 532 (100%) |
| Protocol dosing | 423 (24%) | 303 (100%) | 0 (0%) | 0 (0%) | 120 (100%) | 0 (0%) | 0 (0%) | 0 (0%) | 0 (0%) |
| **Primaquine supervision** |  |  |  |  |  |  |  |  |  |
| Fully supervised | 1,571 (90%) | 303 (100%) | 38 (100%) | 0 (0%) | 120 (100%) | 578 (100%) | 0 (0%) | 0 (0%) | 532 (100%) |
| Partially supervised | 176 (10%) | 0 (0%) | 0 (0%) | 50 (100%) | 0 (0%) | 0 (0%) | 42 (100%) | 84 (100%) | 0 (0%) |
| **Parasite density (asexual parasites/μl)** | 2,515 (617, 7,515) | 760 (320, 2,980) | 2,928 (784, 4,496) | 4,631 (1,993, 8,580) | 872 (144, 2,464) | 4,517 (1,440, 13,565) | 3,471 (1,589, 8,348) | 5,079 (1,913, 12,700) | 2,215 (551, 7,500) |
| **Haemoglobin (g/dl)** | 12.40 (11.30, 13.70) | 11.90 (10.80, 12.80) | 14.20 (13.10, 14.67) | NA (NA, NA) | NA (NA, NA) | 12.50 (11.40, 13.70) | NA (NA, NA) | NA (NA, NA) | 12.65 (11.50, 13.90) |
| Number of missing data | 212 | 0 | 38 | 28 | 120 | 0 | 11 | 15 | 0 |
| **Day-7 methaemoglobin (%)** | 6.0 (3.3, 9.0) | 3.8 (2.6, 5.6) | 5.6 (4.1, 7.5) | 2.5 (1.5, 6.0) | 5.5 (3.5, 8.2) | 6.5 (4.0, 8.9) | 2.4 (1.5, 6.1) | 1.4 (1.2, 7.3) | 7.1 (3.6, 11.0) |
| Number of missing data | 245 | 9 | 3 | 38 | 6 | 68 | 37 | 77 | 7 |
| **Manufacturer of primaquine tablets** | — | Phapros | Shin Poon | Sanofi | Sanofi | Thai Government | Sanofi | Sanofi | Centurion Laboratories |

Numbers are in median (first quartile, third quartile) or frequency (percentage). NA not available, ACT artemisinin-based combination therapy, g gram, dl decilitre, mg milligram, μl microlitre, kg kilogram. § Relapse periodicity was categorised as high (median relapse periodicity of 47 days or less) and low (median relapse periodicity of more than 47) [23]; # Transmission intensity was categorised as low (<1 case per 1000 person-years), moderate (1 case to <10 cases per 1000 person-years), and high (≥10 cases per 1000 person-years) according to subnational malaria incidence estimates for the median year of study enrolment [24]. Missing day 7 methaemoglobin were linearly imputed with methaemoglobin data on ±2 days. Patients with missing haemoglobin or haematocrit data were excluded for the haemolysis sub-dataset.

### Effect of primaquine on methaemoglobin concentrations

There were consistent increases in methaemoglobin concentration from baseline to day 7 during primaquine treatment for both the 7-day and 14-day regimens. On average, the peak observed methaemoglobin concentrations were on day 7 following start of primaquine administration, with a median day 7 methaemoglobin level of 6.0% (IQR: 3.3 to 9.0). Subsequently, methaemoglobin levels tended to decrease slowly in the 7-day or plateau in the 14-day regimen groups (Fig 3, Fig S3).

**Figure 3.**
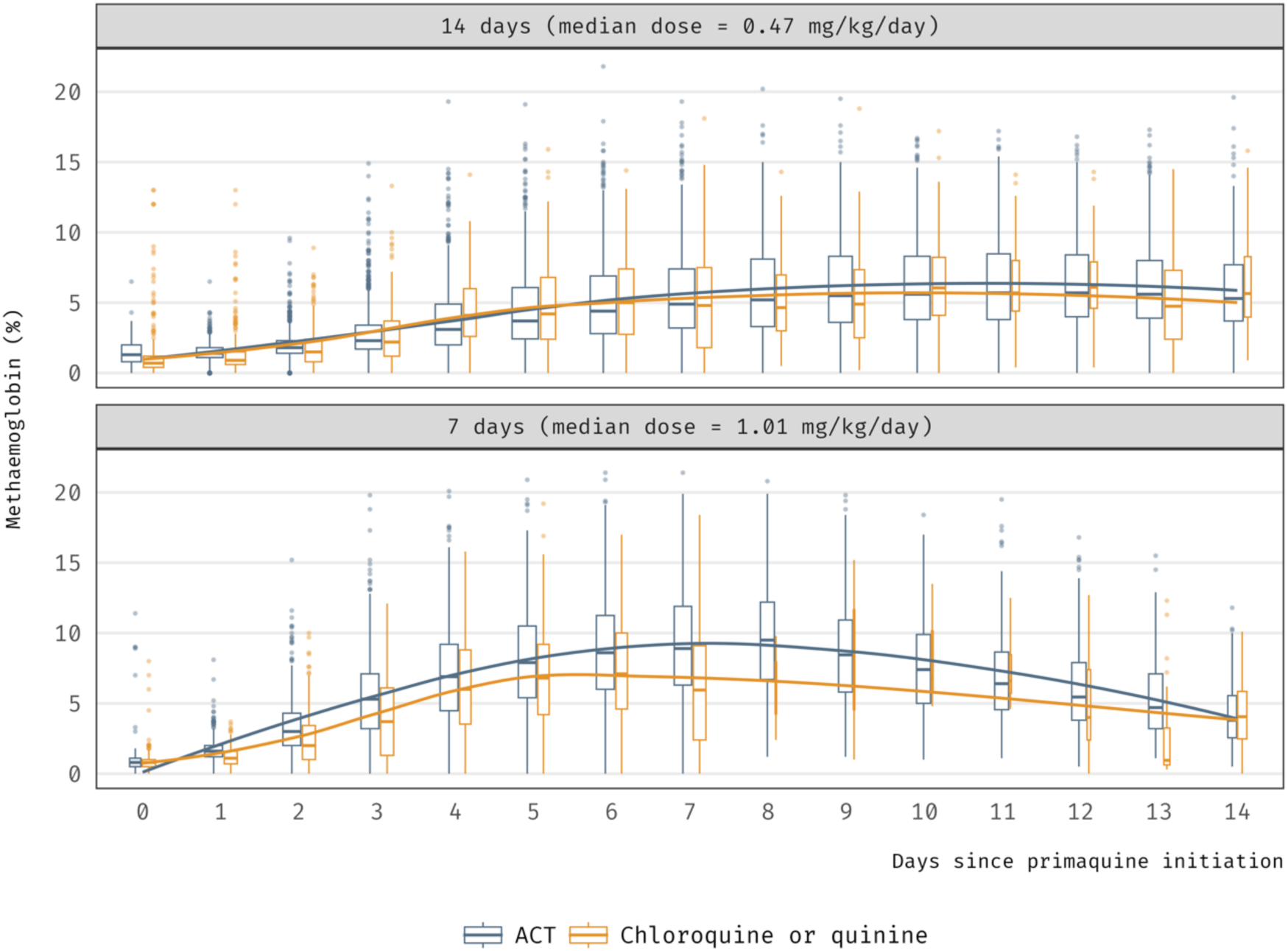
Dynamics of primaquine-induced increases in blood methaemoglobin over time, stratified by primaquine regimen and schizontocidal drug. Methaemoglobin levels increased after starting primaquine in both regimens, usually reaching a maximum after about a week. Methaemoglobin increased at a faster rate among the 7-day regimen reflecting the higher daily dose taken and generally methaemoglobin started to decrease during the second week, when primaquine was no longer administered. Meanwhile, after peaking at also day 7 for the 14-day regimen, methaemoglobin appears to be at a more constant level during the second week. Box width is proportional to the square root of the number of patients. ACT artemisinin-based combination therapy (artesunate/amodiaquine, artesunate/pyronaridine, dihydroartemisinin/piperaquine).

Among patients treated with a low-to-intermediate daily primaquine dose, day 7 methaemoglobin was lower in the studies where primaquine was combined with chloroquine or quinine as a partner drug (Fig S4). However, there was no clear evidence suggesting a drug-drug interaction at the patient level as there was insufficient within-site variation in the pooled data (within-site interaction p = 0.29; across-site interaction p = 0.028). Younger and older patients had lower primaquine-induced methaemoglobinaemia; with the highest levels observed among adolescent patients (Fig S5). Figure S6 shows the distribution of day 7 methaemoglobin concentrations by primaquine regimen.

There was dose-dependent primaquine-induced methaemoglobin production. On average, for every additional 0.1 mg/kg increase in the daily primaquine dose, there was an associated 0.34 percentage-point increase in the day-7 methaemoglobin concentration (95% confidence interval [CI] = [0.16, 0.52]; p = 0.002). This effect remained consistent across all sites (between-site standard deviation in mean difference = 0.20; this quantifies the variability in the estimated effect of the daily primaquine dose on the day-7 methaemoglobin concentration across different sites). For a particular mg/kg daily dose, patients who later developed vivax recurrences (largely attributable to relapses) during follow-up had lower day 7 methaemoglobin values (Fig 4).

**Figure 4.**
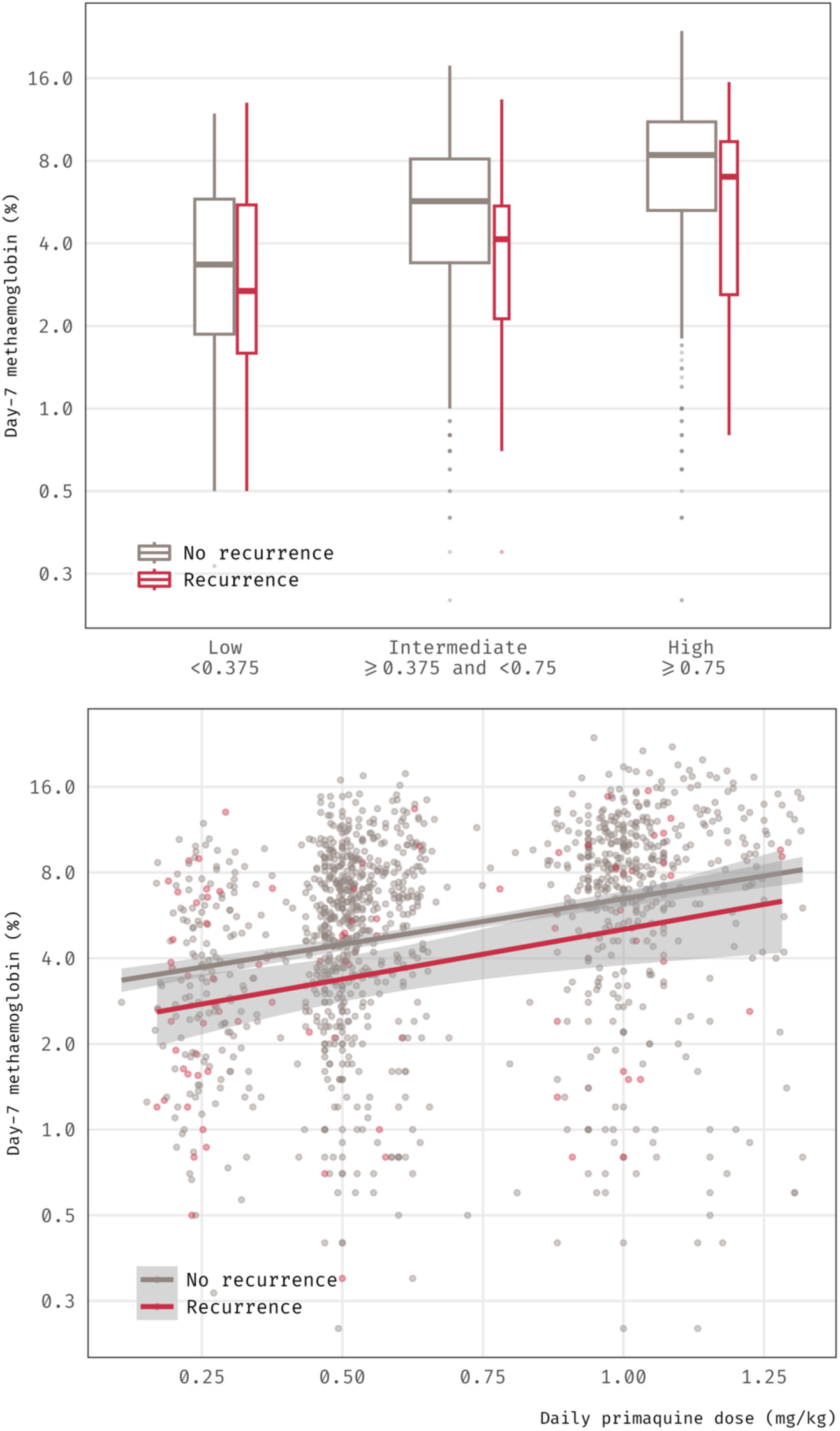
Day 7 blood methaemoglobin (%) as a function of daily mg/kg primaquine dose and *Plasmodium vivax* recurrence status. There was an increasing trend of day 7 methaemoglobin as daily primaquine dose increased among patients with at least 120 days of follow up. Patients who developed *P. vivax* recurrences typically had lower day 7 methaemoglobin levels. Vertical axis is show on the logarithmic scale. Box width is proportional to the square root of the number of patients.

### Association of day 7 methaemoglobin concentrations with the risk of vivax recurrence

After adjusting for the daily and total dose of primaquine and other covariates, a doubling in the observed or imputed day 7 methaemoglobin percentage was associated with an estimated 30% reduction in the risk of vivax recurrence (adjusted hazard ratio [aHR] = 0.70; 95% CI = [0.57, 0.86]; p = 0.0005). These pooled estimates were largely consistent across the study sites (between-site standard deviation in aHR = 1.01; this is a multiplicative factor reflecting the variability in the estimated predictive-effect of the day-7 methaemoglobin concentration on the hazard across different study sites). There was no evidence of proportional hazards violations within the 120 days of follow-up. A sensitivity analysis restricted to patients with observed day 7 methaemoglobin values gave similar estimates (aHR = 0.66; 95% CI = [0.52, 0.84]; p = 0.0008; n = 1502). Controlling primaquine daily dose and duration, the relationship between day 7 methaemoglobin and the risk of vivax recurrence was generally consistent between the studies (Fig 5). A sensitivity analysis using a two-stage approach estimated a slightly smaller effect (dose-adjusted HR for vivax recurrence of 0.81 for each doubling in day 7 methaemoglobin percentage; 95% CI = [0.67, 0.98]; p = 0.037). This numerically different value estimated by the two-stage approach was primarily a result of less flexibility in model specification (e.g., fewer adjustment factors allowed to avoid estimation issues) and sparsity of data.

**Figure 5.**
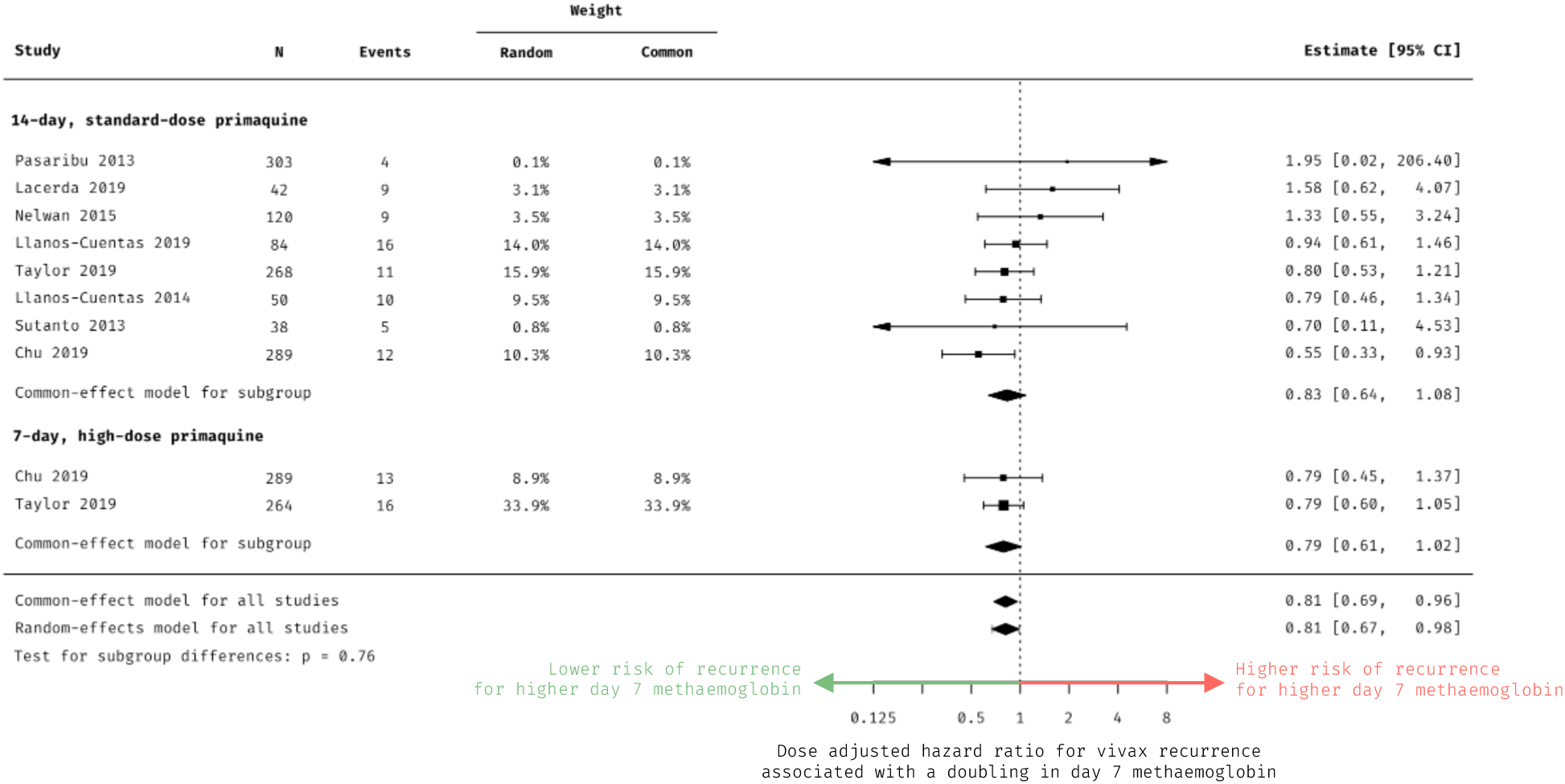
Forest plot. Common-effect and random-effects models converged to comparable estimates. Horizontal axis for dose-adjusted hazard ratios is shown on the logarithmic scale.

On the absolute scale (i.e. percentage of the total haemoglobin), each additional percentage-point increase in day 7 methaemoglobin was associated with an estimated 10% reduction in the risk of vivax recurrence (aHR = 0.90; 95% CI = [0.84, 0.96]; p=0.003). Improved model-fit (p<0.0001) was observed by specifying day 7 methaemoglobinaemia on the logarithmic scale (as in the main model), indicating linearity between multiplicative changes in day-7 methaemoglobin level and the log-hazard ratio for vivax recurrence.

### Additional analyses

Of the 1344 (77%) patients followed for at least 120 days, we observed a comparable relationship between day 7 methaemoglobin and the risk of any observed vivax recurrence (dose-adjusted odds ratio of 0.66 for each doubling in day 7 methaemoglobin; 95% CI = [0.52, 0.83]; p=0.0004). Of the 1360 (78%) G6PD normal patients with haemoglobin (or haematocrit) measured at baseline, there was little or no evidence of an association between the maximum absolute decrease in haemoglobin concentration from day 0 to days 2–3 and the day-7 methaemoglobinaemia (adjusted mean difference = 0.01; 95% CI = [–0.21, 0.23] p = 0.90).

Assuming a comparison between two primaquine doses whereby the higher dose results in half the risk of vivax recurrence with recurrence rates of 16% versus 8% (corresponding to primaquine doses of 0.5 versus 1 mg/kg over 7 days), a two-arm randomised trial aiming to show superiority would require a sample size of 256 individuals per group to achieve 80% power with a 5% false positive rate. In contrast, using day 7 methaemoglobin as a surrogate endpoint for vivax recurrence, the required sample size would be reduced by approximately 42% (n = 148 individuals per group) for high daily dose primaquine and could be even more for lower daily doses (Fig S7).

## Discussion

In this systematic review and individual patient data meta-analysis, we confirm that higher primaquine-induced methaemoglobin concentrations on day 7 are associated with lower rates of *P. vivax* recurrence. This finding remained consistent across diverse study sites with varying populations and levels of transmission intensity. Our analysis found no indication of a differential predictive effect of day 7 methaemoglobin between the 7-day and 14-day primaquine regimens. Additionally, we observed a positive dose-response relationship between the daily weight-adjusted primaquine dose and day 7 methaemoglobin level.

Our findings are in line with recent estimates derived from Chu 2021 [10] (a secondary analysis of primaquine trial [9] data from the endemic northwest Thailand-Myanmar border) and Watson 2022 [4] (an individual patient data meta-analysis of tafenoquine trials). These estimates suggested that an increase of one percentage-point of day 7 methaemoglobinaemia was associated with an estimated reduction of 10% for primaquine and 20% for tafenoquine in the risk of *P. vivax* recurrence. While our analysis of primaquine included data from these studies such that estimates are not completely uncorrelated, additional data from different studies contributed most (nearly 70%) of our pooled dataset. Watson 2022 analysed vivax patients who received tafenoquine only (not included in this analysis). According to the White 2022 review [8] of early experiments with primaquine or its analogues conducted more than 70 years ago [30–32], 8-aminoquinoline analogues which resulted in less than 6% of methaemoglobinaemia during treatment showed reduced efficacy. All these results point towards higher methaemoglobin levels following treatment as indicative of increased antihypnozoite activity.

In the current analysis of G6PD normal patients, we observed no evidence of an association between the extent of early haemolysis on days 2–3 from baseline and day 7 methaemoglobinaemia. This suggests that during the early days of illness, early haemolysis was primarily attributable to the acute parasitaemia and rehydration, rather than iatrogenic haemolysis caused by the primaquine active metabolites. The lower day 7 methaemoglobin observed among younger patients may be partly explained by age-related enzyme immaturity and lower drug exposures [10]. The reason for lower methaemoglobin in older patients is less clear.

Methaemoglobinaemia is a simple and readily measured surrogate endpoint that can be quantified within a week after starting administration of an 8-aminoquinoline and has potential to improve the efficiency of exploratory trials (drug-drug interactions, regimen optimisation, drug screening, dose optimisation). Current antirelapse clinical trials generally require patient follow up for many months to observe recurrent infections in order to have sufficient power to determine comparative efficacy of different treatment arms. Quantifying methaemoglobinaemia also has potential to be a useful approach for monitoring of trials and clinical practices at an individual level, serving as a surrogate marker for patient adherence. Although methaemoglobin is a surrogate endpoint for adherence to treatment, these results are not driven by adherence (90% of patients had fully supervised treatment). Early-phase studies, preceding more definitive efficacy trials, may stand to benefit the most from this novel endpoint [8]. Minimum required sample sizes could also be substantially reduced by using day 7 methaemoglobin as an endpoint to improve cost-efficiency in conducting *P. vivax* trials. An ongoing study (NCT05788094) is currently using day 7 methaemoglobin as a secondary outcome to address the question of whether there is important drug-drug interaction between tafenoquine and chloroquine, DHA-piperaquine, or artemether-lumefantrine [33]. The recent INSPECTOR trial in Indonesia used DHA-piperaquine as the partner schizonticidal drug and lower than expected efficacy was observed [34] compared to trials with chloroquine [35–37]. In the INSPECTOR study, tafenoquine plus DHA-piperaquine resulted in a median methaemoglobin of 1.3% (range = 0.7% to 3.7%), which was similar to DHA-piperaquine alone (median = 1.0%, range = 0.5% to 1.8%). In contrast, primaquine plus DHA-piperaquine produced more than twice as much methaemoglobin (median = 2.9%, range = 0.9% to 7.9%) and superior efficacy than the tafenoquine plus DHA-piperaquine arm.

Our study has several limitations. Some patients missed measurements and some studies by design intentionally did not measure methaemoglobin on day 7. We addressed such missing data through linear interpolation, since methaemoglobin was recorded within two days before and/or after day 7. It is important to note that methaemoglobinaemia itself is an inherently noisy measure with substantial variation between patients, especially in patients receiving higher doses of 8-aminoquinoline treatment (Fig S7 shows higher methaemoglobin variability in higher dose groups). Improved reliability may be achieved by measuring methaemoglobin on both hands, incorporating multiple or repeated measurements, waiting longer, and blocking fluorescent light (e.g., turning off lights or covering hand when checking). In an upcoming analysis, we plan to explore alternative summary metrics for methaemoglobin levels using pharmacometric modelling [22]. This approach aims to capture exposures to primaquine’s active metabolites and improve the overall robustness of our findings. There is also a possibility of less accurate methaemoglobin measurements by using CO-oximetry for patients with dark skin pigment [38].

Only two studies [39, 40] were conducted in locations with negligible reinfection. Hence another limitation of our analysis is the uncertainty regarding the aetiology of recurrences for most of the patients, since there is currently no standardised method to differentiate between recrudescence, reinfection, and relapse [1]. It is important to note that the use of highly efficacious schizonticides across the study sites makes recrudescence unlikely. Another potential source of error includes participants who might not have had hypnozoites in the liver to start with, thus having zero risk of relapse, despite primaquine inducing methaemoglobinaemia. However, this circumstance should bias the effect estimates towards the null (i.e., no association between day 7 methaemoglobinaemia and the risk of vivax recurrence), making our estimates conservative.

The generalisability of our findings is constrained to the large majority of vivax malaria patients with normal G6PD activity (≥30% G6PD activity or a negative qualitative test). For individuals with G6PD deficiency, blood methaemoglobin will not serve as a valid surrogate endpoint for antihypnozoite activity [8, 10]. In G6PD deficiency, primaquine does not induce clear methaemoglobinaemia [15, 41]. We currently lack sufficient data to explore how *CYP2D6* polymorphism impacts primaquine biotransformation to its active metabolites, as most patients were not genotyped [10]. The previous analysis by Chu et al suggested lower methaemoglobin in null *CYP2D6* metabolisers (activity score of 0) [8]. Intermediate metabolisers may also have lower active metabolites and higher relapse rates [6, 7]. Further research specifically targeted at these vulnerable populations is imperative.

Our pooled dataset includes patients from various countries with different antimalarial treatment policies for their first-line schizonticides, each having different elimination kinetics and thus different durations of suppressive post-treatment prophylaxis. The use of time-to-event models in such contexts may introduce bias in favour of drugs with longer half-lives, such as chloroquine. However, our sensitivity analysis, employing logistic regression that included partner drug as a covariate, yielded similar predictive-effect estimates.

In conclusion, in primaquine-treated G6PD normal individuals, the day 7 methaemoglobin can serve as a pharmacodynamic proxy for exposure to the biologically active metabolites of primaquine, making it a valid surrogate endpoint in G6PD normal patients with *P. vivax* malaria. The consistency of results observed in tafenoquine studies suggests a common drug-class phenomenon for 8-aminoquinolines. Direct comparisons of 8-aminoquinoline induced methaemoglobinaemia between primaquine and tafenoquine in future studies could prove useful. These findings collectively enhance our understanding of the causal mechanisms by which 8-aminoquinoline drugs exert their effects, facilitate drug discovery and regimen optimisation, and influence clinical practices.

## Data Availability

Pseudonymised participant data used in this study can be accessed via the WorldWide Antimalarial Resistance Network (wwarn.org). Requests for access will be reviewed by a data access committee to ensure that use of data protects the interests of the participants and researchers according to the terms of ethics approval and principles of equitable data sharing. Requests can be submitted by email to via the data access form available at https://www.wwarn.org/working-together/sharing-accessing-data/accessing-data. WWARN is registered with the Registry of Research Data Repositories (https://www.re3data.org/).

## Acknowledgements

We thank all patient volunteers, healthcare workers, and research staff who contributed to the individual studies. We thank the WWARN team for technical and administrative support. IF is supported by the Oxford Nuffield Department of Medicine Tropical Network Fund. RNP and RJC are supported by Australian National Health and Medical Research Council (NHMRC) Investigator Grants (2008501 and 1194701, respectively). Shoklo Malaria Research Unit (grant 220211) and FN (grant 089179) are supported by the Wellcome Trust. MVGL is a fellow from the National Council for Scientific and Technological Development (CNPq). KT is a CSL Centenary Fellow. JAW is a Sir Henry Dale Fellow funded by the Wellcome Trust (223253/Z/21/Z). NJW is a Wellcome Trust Principal Fellow (093956/Z/10/C). This research was supported grants from the Wellcome Trust. For the purpose of open access, the authors have applied a CC BY public copyright licence to any Author Accepted Manuscript arising from this submission.

## Author contributions

IF, RJC, RNP, NJW, JKB, and JAW conceived the study, analysed and interpreted the data, as well as drafted the manuscript. IF, RJC, and JAW accessed and verified the data. NHC, NPJD, JAG, GCKWK, MVGL, AL, EJN, FN, APP, IS, WRJT, KT, RNP, NJW, JKB conceived and undertook the individual studies and enrolled the patients. All authors critically reviewed the study for intellectual content, revised the manuscript, and were responsible for the decision to submit for publication.

## Data sharing

Pseudonymised participant data used in this study can be accessed via the WorldWide Antimalarial Resistance Network (wwarn.org). Requests for access will be reviewed by a data access committee to ensure that use of data protects the interests of the participants and researchers according to the terms of ethics approval and principles of equitable data sharing. Requests can be submitted by email to via the data access form available at https://www.wwarn.org/working-together/sharing-accessing-data/accessing-data. WWARN is registered with the Registry of Research Data Repositories (https://www.re3data.org/). Code for data analysis and visualisation is available at https://github.com/ihsanfadil/methb7.

## Supporting Information

**List S1. Systematic search terms for the databases**

Vivax AND (artefenomel OR arterolane OR amodiaquine OR atovaquone OR artemisinin OR arteether OR artesunate OR artemether OR artemotil OR azithromycin OR artekin OR chloroquine OR chlorproguanil OR cycloguanil OR clindamycin OR coartem OR dapsone OR dihydroartemisinin OR duo-cotecxin OR doxycycline OR halofantrine OR lumefantrine OR lariam OR malarone OR mefloquine OR naphthoquine OR naphthoquinone OR piperaquine OR primaquine OR proguanil OR pyrimethamine OR pyronaridine OR proguanil OR quinidine OR quinine OR riamet OR sulphadoxine OR tetracycline OR tafenoquine)

**Table S1.** PRISMA-IPD checklist.

| PRISMA-IPD<br>Section/topic | Item<br>No | Checklist item | Reported<br>on page |
| --- | --- | --- | --- |
| Title |  |  |  |
| Title | 1 | Identify the report as a systematic review and meta-analysis of individual participant data. | 1 |
| Abstract |  |  |  |
| Structured<br>summary | 2 | Provide a structured summary including as applicable: | 2 |
|  |  | **Background:** state research question and main objectives, with information on participants, interventions, comparators and outcomes. |  |
|  |  | **Methods:** report eligibility criteria; data sources including dates of last bibliographic search or elicitation, noting that IPD were sought; methods of assessing risk of bias. |  |
|  |  | **Results:** provide number and type of studies and participants identified and number (%) obtained; summary effect estimates for main outcomes (benefits and harms) with confidence intervals and measures of statistical heterogeneity. Describe the direction and size of summary effects in terms meaningful to those who would put findings into practice. |  |
|  |  | **Discussion:** state main strengths and limitations of the evidence, general interpretation of the results and any important implications. |  |
|  |  | **Other:** report primary funding source, registration number and registry name for the systematic review and IPD meta-analysis. |  |
| Introduction |  |  |  |
| Rationale | 3 | Describe the rationale for the review in the context of what is already known. | 3 |
| Objectives | 4 | Provide an explicit statement of the questions being addressed with reference, as applicable, to participants, interventions, comparisons, outcomes and study design (PICOS). Include any hypotheses that relate to particular types of participant-level subgroups. | 3 |
| Methods |  |  |  |
| Protocol and registration | 5 | Indicate if a protocol exists and where it can be accessed. If available, provide registration information including registration number and registry name. Provide publication details, if applicable. | 4 |
| Eligibility criteria | 6 | Specify inclusion and exclusion criteria including those relating to participants, interventions, comparisons, outcomes, study design and characteristics (e.g. years when conducted, required minimum follow-up). Note whether these were applied at the study or individual level i.e. whether eligible participants were included (and ineligible participants excluded) from a study that included a wider population than specified by the review inclusion criteria. The rationale for criteria should be stated. | 4 |
| Identifying studies - | 7 | Describe all methods of identifying published and unpublished studies including, as applicable: which bibliographic databases were searched with dates of coverage; details of any hand searching including of conference proceedings; use of study registers | 4 |
| information sources |  | and agency or company databases; contact with the original research team and experts in the field; open adverts and surveys. Give the date of last search or elicitation. |  |
| Identifying studies - search | 8 | Present the full electronic search strategy for at least one database, including any limits used, such that it could be repeated. | 24 |
| Study selection processes | 9 | State the process for determining which studies were eligible for inclusion. | 4 |
| Data collection processes | 10 | Describe how IPD were requested, collected and managed, including any processes for querying and confirming data with investigators. If IPD were not sought from any eligible study, the reason for this should be stated (for each such study). | 4 |
|  |  | If applicable, describe how any studies for which IPD were not available were dealt with. This should include whether, how and what aggregate data were sought or extracted from study reports and publications (such as extracting data independently in duplicate) and any processes for obtaining and confirming these data with investigators. |  |
| Data items | 11 | Describe how the information and variables to be collected were chosen. List and define all study level and participant level data that were sought, including baseline and follow-up information. If applicable, describe methods of standardising or translating variables within the IPD datasets to ensure common scales or measurements across studies. | 4 |
| IPD integrity | A1 | Describe what aspects of IPD were subject to data checking (such as sequence generation, data consistency and completeness, baseline imbalance) and how this was done. | 4–6 |
| Risk of bias assessment in individual studies. | 12 | Describe methods used to assess risk of bias in the individual studies and whether this was applied separately for each outcome. If applicable, describe how findings of IPD checking were used to inform the assessment. Report if and how risk of bias assessment was used in any data synthesis. | 6 |
| Specification of outcomes and effect measures | 13 | State all treatment comparisons of interests. State all outcomes addressed and define them in detail. State whether they were pre-specified for the review and, if applicable, whether they were primary/main or secondary/additional outcomes. Give the principal measures of effect (such as risk ratio, hazard ratio, difference in means) used for each outcome. | 4–6 |
| Synthesis methods | 14 | Describe the meta-analysis methods used to synthesise IPD. Specify any statistical methods and models used. Issues should include (but are not restricted to):<br>• Use of a one-stage or two-stage approach.<br>• How effect estimates were generated separately within each study and combined across studies (where applicable).<br>• Specification of one-stage models (where applicable) including how clustering of patients within studies was accounted for.<br>• Use of fixed or random effects models and any other model assumptions, such as proportional hazards.<br>• How (summary) survival curves were generated (where applicable).<br>• Methods for quantifying statistical heterogeneity (such as $I^2$ and $t^2$). | 4–6 |
|  |  | How studies providing IPD and not providing IPD were analysed together (where applicable).<br>How missing data within the IPD were dealt with (where applicable). |  |
| Exploration of variation in effects | A2 | If applicable, describe any methods used to explore variation in effects by study or participant level characteristics (such as estimation of interactions between effect and covariates). State all participant-level characteristics that were analysed as potential effect modifiers, and whether these were pre-specified. | 5–6 |
| Risk of bias across studies | 15 | Specify any assessment of risk of bias relating to the accumulated body of evidence, including any pertaining to not obtaining IPD for particular studies, outcomes or other variables. | 6 |
| Additional analyses | 16 | Describe methods of any additional analyses, including sensitivity analyses. State which of these were pre-specified. | 5–6 |
| **Results** |  |  |  |
| Study selection and IPD obtained | 17 | Give numbers of studies screened, assessed for eligibility, and included in the systematic review with reasons for exclusions at each stage. Indicate the number of studies and participants for which IPD were sought and for which IPD were obtained. For those studies where IPD were not available, give the numbers of studies and participants for which aggregate data were available. Report reasons for non-availability of IPD. Include a flow diagram. | 6, 7, 18 |
| Study characteristics | 18 | For each study, present information on key study and participant characteristics (such as description of interventions, numbers of participants, demographic data, unavailability of outcomes, funding source, and if applicable duration of follow-up). Provide (main) citations for each study. Where applicable, also report similar study characteristics for any studies not providing IPD. | 6, 7, 19, 20, 32–35 |
| IPD integrity | A3 | Report any important issues identified in checking IPD or state that there were none. | 7, 34 |
| Risk of bias within studies | 19 | Present data on risk of bias assessments. If applicable, describe whether data checking led to the up-weighting or down-weighting of these assessments. Consider how any potential bias impacts on the robustness of meta-analysis conclusions. | 6, 7, 30 |
| Results of individual studies | 20 | For each comparison and for each main outcome (benefit or harm), for each individual study report the number of eligible participants for which data were obtained and show simple summary data for each intervention group (including, where applicable, the number of events), effect estimates and confidence intervals. These may be tabulated or included on a forest plot. | 23 |
| Results of syntheses | 21 | Present summary effects for each meta-analysis undertaken, including confidence intervals and measures of statistical heterogeneity. State whether the analysis was pre-specified, and report the numbers of studies and participants and, where applicable, the number of events on which it is based. | 8, 9 |
|  |  | When exploring variation in effects due to patient or study characteristics, present summary interaction estimates for each characteristic examined, including confidence intervals and measures of statistical heterogeneity. State whether the analysis was pre-specified. State whether any interaction is consistent across trials. |  |
|  |  | Provide a description of the direction and size of effect in terms meaningful to those who would put findings into practice. |  |
| Risk of bias across studies | 22 | Present results of any assessment of risk of bias relating to the accumulated body of evidence, including any pertaining to the availability and representativeness of available studies, outcomes or other variables. | 30 |
| Additional analyses | 23 | Give results of any additional analyses (e.g. sensitivity analyses). If applicable, this should also include any analyses that incorporate aggregate data for studies that do not have IPD. If applicable, summarise the main meta-analysis results following the inclusion or exclusion of studies for which IPD were not available. | 8, 9 |
| **Discussion** |  |  |  |
| Summary of evidence | 24 | Summarise the main findings, including the strength of evidence for each main outcome. | 9 |
| Strengths and limitations | 25 | Discuss any important strengths and limitations of the evidence including the benefits of access to IPD and any limitations arising from IPD that were not available. | 10, 11 |
| Conclusions | 26 | Provide a general interpretation of the findings in the context of other evidence. | 9, 10 |
| Implications | A4 | Consider relevance to key groups (such as policy makers, service providers and service users). Consider implications for future research. | 11 |
| **Funding** |  |  |  |
| Funding | 27 | Describe sources of funding and other support (such as supply of IPD), and the role in the systematic review of those providing such support. | 11, 12 |

**List S2. Signalling questions for risk of bias assessment using the QUIPS tool adapted to the current analysis**

<u>Domain 1: The study sample represents the population of interest on key characteristics, sufficient to limit potential bias of the observed relationship between the predictive factor and outcome.</u>

- The source population or population of interest is adequately described.
- The baseline study sample (i.e., individuals entering the study) is adequately described.
- The sampling frame and recruitment are adequately described, including methods to identify the sample sufficient to limit potential bias.
- Period of recruitment is adequately described.
- Place of recruitment (setting, level of endemicity, geographic location) are adequately described.
- Inclusion and exclusion criteria are adequately described).

<u>Domain 2: Loss to follow up (from baseline sample to study population analysed) is not associated with certain characteristics (i.e., the study data adequately represent the sample) sufficient to limit potential bias to the observed relationship between predictive factor and outcome.</u>

- Response rate (i.e., proportion of study sample completing the study and providing outcome data) is adequate.
- Attempts to collect information on participants who dropped out of the study are described.
- Reasons for loss to follow up are provided.
- Participants lost to follow up are adequately described.
- There are no important differences between participants who completed the study and those who did not.

<u>Domain 3: Predictive factor and drug intervention are adequately measured in study participants to sufficiently limit potential bias.</u>

- A clear definition or description of the primaquine regimen and measured methaemoglobin is provided (e.g., including dose, level, duration of exposure, and clear specification of the method of measurement).
- Adequately accurate and reliable measurement of primaquine doses and methaemoglobin concentrations to limit misclassification bias.
- Continuous variables are reported, or clinically relevant cut points (i.e., not data-dependent) are used.
- Method and setting of methaemoglobin measurement are the same for all study participants.
- Adequate proportion of the study sample has complete methaemoglobin data.
- Adequate adherence or supervision of primaquine administration.

<u>Domain 4: Outcome of interest is adequately measured in study participants to sufficiently limit potential bias.</u>

- A clear definition of the outcome is provided, including duration of follow up.
- Method of outcome measurement used is adequately accurate and reliable to limit misclassification bias.
- Method and setting of outcome measurement are the same for all study participants.

**Table S2.**
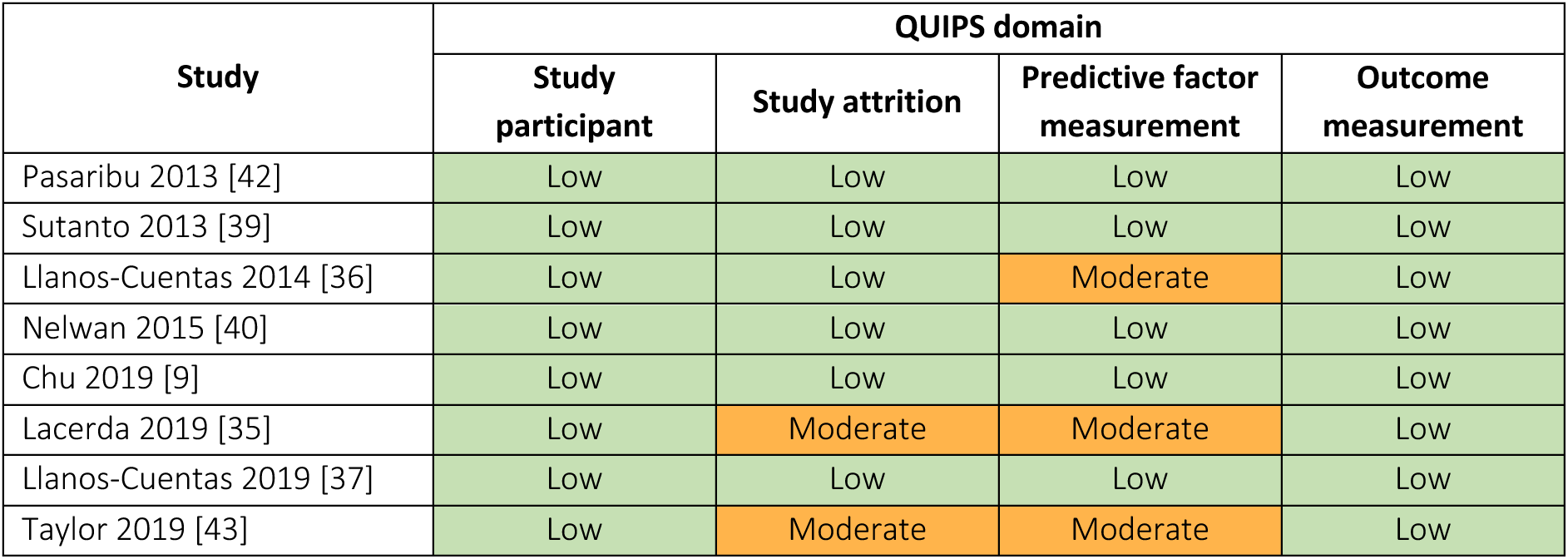
Risk of bias assessment.

**Figure S1.**
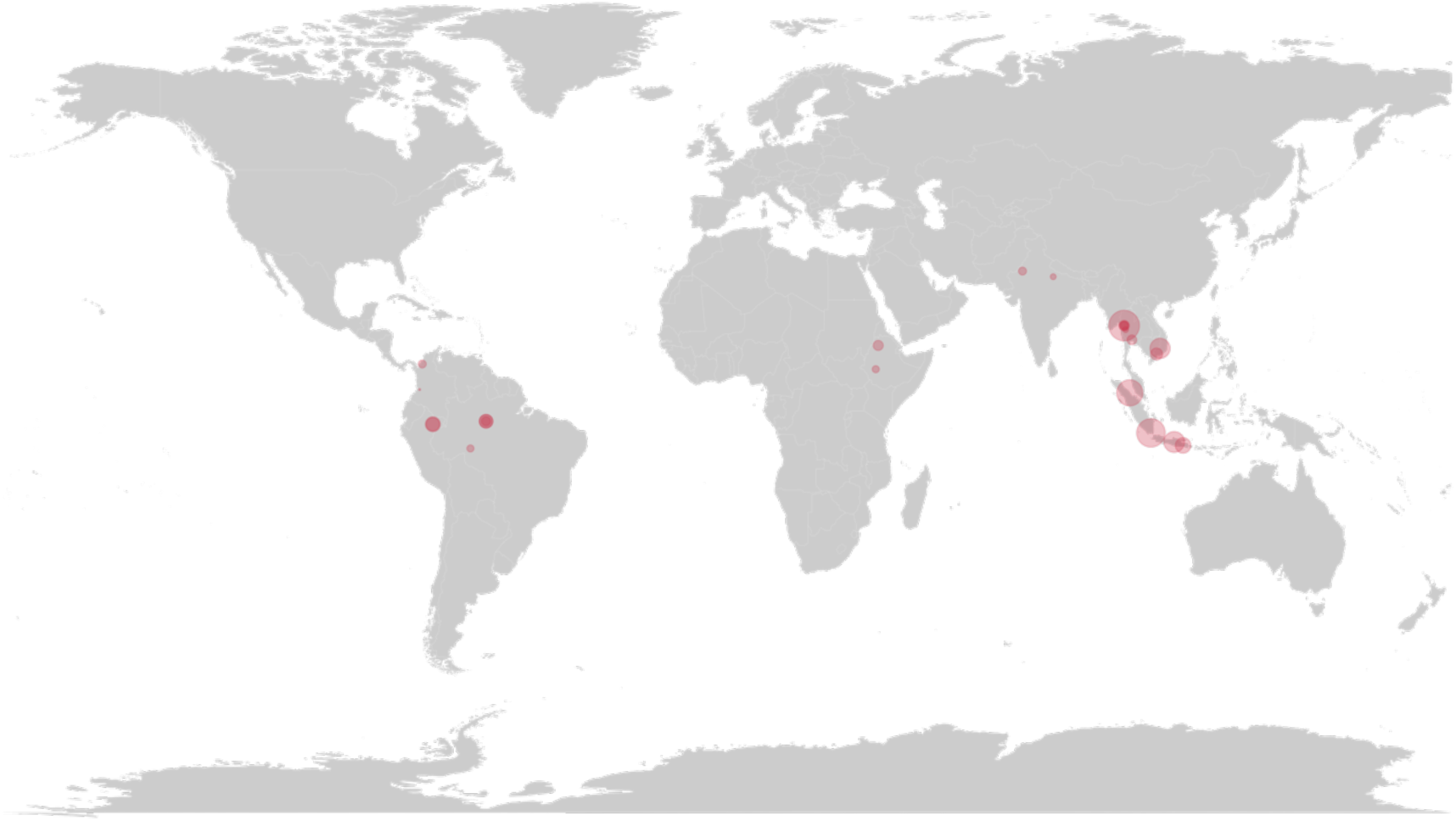
Study sites that contributed to the pooled data in this individual patient data meta-analysis. Red bubble represents a study site with size proportional to the square root of the number of patients.

**Table S3.** Studies included in analysis.

| Paper | Study site | Country | Region | Latitude | Longitude | Year start | Year end | MAP incidence rate (per 1000 persons) | Transmission intensity# | Relapse periodicity§ |
| --- | --- | --- | --- | --- | --- | --- | --- | --- | --- | --- |
| **Pasaribu 2013 [42]** | Tanjung Leidong | Indonesia | Asia-Pacific | 2.77 | 99.98 | 2011 | 2011 | 2.75 | Moderate | High |
| **Sutanto 2013 [39]** | Lumajang | Indonesia | Asia-Pacific | -8.13 | 113.22 | 2010 | 2011 | 36.88* | High | High |
| **Llanos-Cuentas 2014 [36]** | Bangkok | Thailand | Asia-Pacific | 13.76 | 100.50 | 2011 | 2013 | 0.16 | Low | High |
| **Llanos-Cuentas 2014 [36]** | Mae Sot | Thailand | Asia-Pacific | 16.72 | 98.58 | 2011 | 2013 | 3.07 | Moderate | High |
| **Llanos-Cuentas 2014 [36]** | Lucknow | India | Asia-Pacific | 26.85 | 80.95 | 2011 | 2013 | 2.84 | Moderate | Low |
| **Llanos-Cuentas 2014 [36]** | Bikaner | India | Asia-Pacific | 28.02 | 73.31 | 2011 | 2013 | 2.85 | Moderate | Low |
| **Llanos-Cuentas 2014 [36]** | Iquitos | Peru | Americas | -3.74 | -73.25 | 2011 | 2013 | 40.49 | High | Low |
| **Llanos-Cuentas 2014 [36]** | Manaus | Brazil | Americas | -3.12 | -60.02 | 2011 | 2013 | 42.81 | High | Low |
| **Nelwan 2015 [40]** | Sragen | Indonesia | Asia-Pacific | -7.42 | 111.02 | 2013 | 2013 | 42.44* | High | High |
| **Chu 2019 [9]** | Mae Sot | Thailand | Asia-Pacific | 16.72 | 98.58 | 2012 | 2014 | 3.09 | Moderate | High |
| **Lacerda 2019 [35]** | Manaus | Brazil | Americas | -3.12 | -60.02 | 2013 | 2016 | 41.14 | High | Low |
| **Lacerda 2019 [35]** | Porto Velho | Brazil | Americas | -8.76 | -63.90 | 2013 | 2016 | 9.23 | Moderate | Low |
| **Lacerda 2019 [35]** | Jimma | Ethiopia | Africa | 7.67 | 36.84 | 2013 | 2016 | 40.53 | High | Low |
| **Lacerda 2019 [35]** | Gondar | Ethiopia | Africa | 12.60 | 37.45 | 2013 | 2016 | 6.72 | Moderate | Low |
| **Lacerda 2019 [35]** | Mae Sot | Thailand | Asia-Pacific | 16.72 | 98.58 | 2013 | 2016 | 0.64 | Low | High |
| **Llanos-Cuentas 2019 [37]** | Manaus | Brazil | Americas | -3.12 | -60.02 | 2015 | 2016 | 18.55 | High | Low |
| **Llanos-Cuentas 2019 [37]** | Monteira | Colombia | Americas | 8.75 | -75.88 | 2015 | 2016 | 5.36 | Moderate | Low |
| **Llanos-Cuentas 2019 [37]** | Cali | Colombia | Americas | 3.45 | -76.53 | 2015 | 2016 | 1.93 | Moderate | Low |
| **Llanos-Cuentas 2019 [37]** | Iquitos | Peru | Americas | -3.74 | -73.25 | 2015 | 2016 | 58.05 | High | Low |
| **Llanos-Cuentas 2019 [37]** | Umphang | Thailand | Asia-Pacific | 15.88 | 98.92 | 2015 | 2016 | 1.08 | Moderate | High |
| **Llanos-Cuentas 2019 [37]** | Mae Sot | Thailand | Asia-Pacific | 16.72 | 98.58 | 2015 | 2016 | 1.08 | Moderate | High |
| **Llanos-Cuentas 2019 [37]** | Ho Chi Minh City | Vietnam | Asia-Pacific | 10.82 | 106.63 | 2015 | 2016 | 0.01 | Low | High |
| **Taylor 2019 [43]** | Dak O | Vietnam | Asia-Pacific | 12.00 | 107.50 | 2015 | 2017 | 0.24 | Low | High |
| **Taylor 2019 [43]** | Hanura | Indonesia | Asia-Pacific | -5.53 | 105.24 | 2015 | 2017 | 1.01 | Moderate | High |

MAP malaria atlas project. § Relapse periodicity was categorised as high (median relapse periodicity of 47 days or less) and low (median relapse periodicity of more than 47) [23]; # Transmission intensity was categorised as low (<1 case per 1000 person-years), moderate (1 case to <10 cases per 1000 person-years), and high (≥10 cases per 1000 person-years) according to subnational malaria incidence estimates for the median year of study enrolment [24]. \*Based on the location where patients were infected by *P. vivax* in Indonesian Papua.

**Figure S2.**
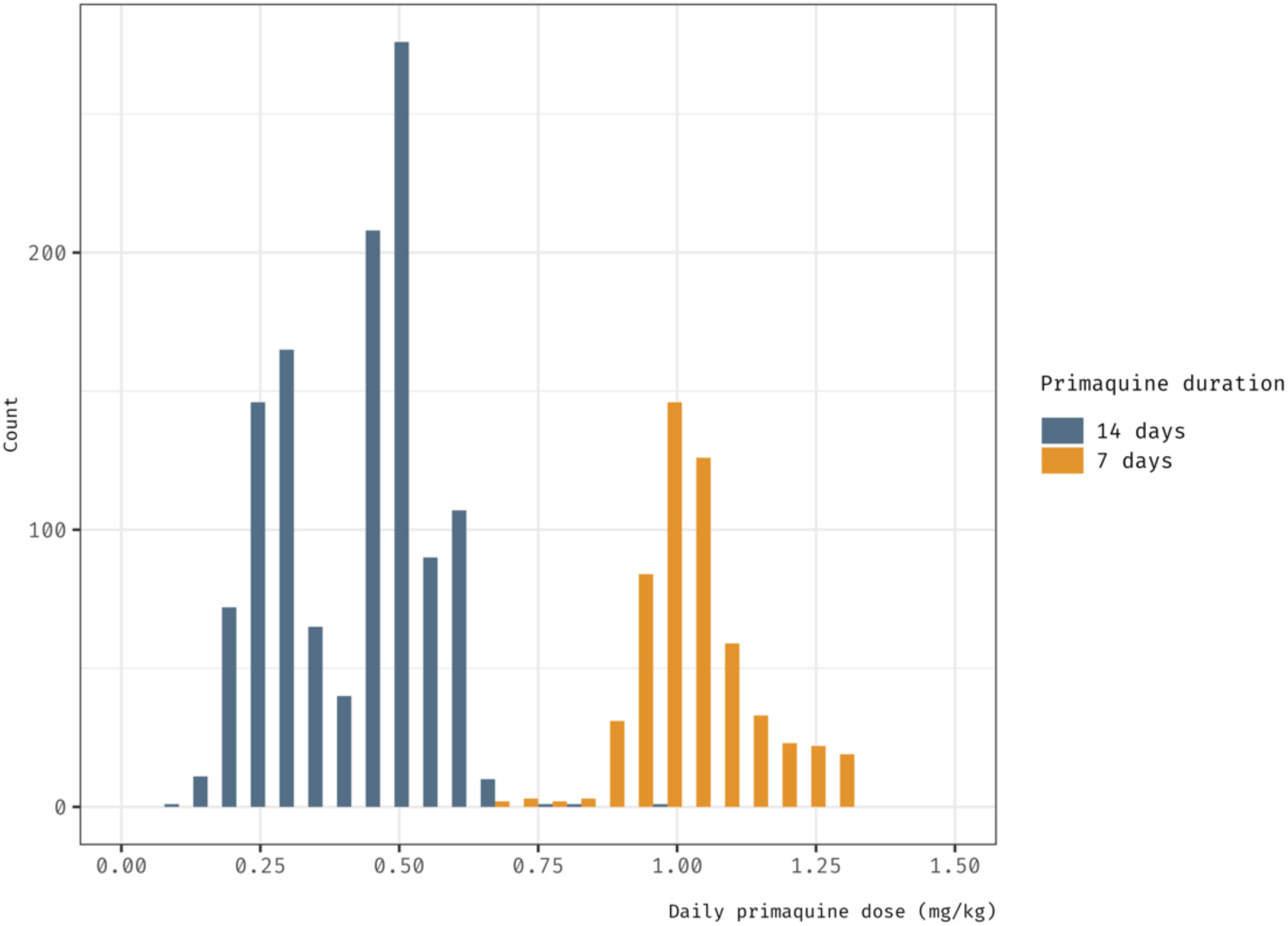
Distribution of weight-adjusted primaquine daily dose by primaquine regimen. In the 14-day primaquine regimen, the observed two peaks reflect the targeted total primaquine dose of 3.5 and 7 mg (base) per kg body weight.

**Table S4.** Studies that were eligible for analysis but not included in the pooled data.

| Characteristic | Study |  |  |  |  |
| --- | --- | --- | --- | --- | --- |
|  | Solari Soto [29] | Carmona-Fonseca [25] | Ley [28] | Fukuda [27] | Chu [26] |
| **Year published** | 2002 | 2010 | 2016 | 2017 | 2018 |
| **Number of treatment arms** | 2 | 2 | 1 | 2 | 3 |
| **Number of sites** | 1 | 1 | 2 | 1 | 1 |
| **Region** | Americas | Americas | Asia-Pacific | Asia-Pacific | Asia-Pacific |
| **Country** | Peru | Colombia | Bangladesh | Thailand | Thailand |
| **Follow-up (days)** | 60 | 120 | 28 | 120 | 365 |
| **Randomised** | Yes | Yes | No | Yes | Yes |
| **Recruitment period** | 1998–1999 | 2005–2008 | 2014–2015 | 2003–2005 | 2010–2012 |
| **Treatment arms** | (1)<br>Chloroquine + 14-day, low-dose primaquine,<br>(2)<br>Chloroquine + 7-day, low-dose primaquine | (1)<br>Chloroquine + 7-day, low-dose primaquine,<br>(2)<br>Chloroquine + 3-day, low-dose primaquine | Chloroquine + 14-day, low-dose primaquine | (1)<br>Chloroquine + 14-day, low-dose primaquine,<br>(2)<br>Tafenoquine | (1) Artesunate,<br>(2)<br>Chloroquine,<br>(3)<br>Chloroquine + 14-day, high-dose primaquine |
| ***P. vivax* patients enrolled** | 60 | 79 | 66 | 70 | 644 |
| **Treated with primaquine** | 60 | 79 | 66 | 24 | 198 |
| **Supervision** | Yes | Yes | Yes | Yes | Yes |
| **Sex (primaquine receiving arm)** | 57% male | Mostly male | 62% male | 83% male | 64% male |
| **Age (in years, primaquine receiving arm)** | Average = 26.5 | Range = 10–17 | Median = 18 (mono-infection), 14 (mixed) | Median = 30 | Median = 18 |
| **Reason for exclusion** | No response from investigators | Data not available | Missing minimum data | Data not provided | Missing minimum data |

IPD individual patient data.

**Table S5.** Comparison of characteristics of patients (as originally reported) who received primaquine between included and eligible but not included studies.

| Characteristic | Included studies (n = 8) | Eligible but not included studies (n = 5) |
| --- | --- | --- |
| **Region, studies (percentage)** |  |  |
| Asia-Pacific | 21 (58.3%)<sup>€</sup> | 3 (60%) |
| Africa | 4 (11.1%)<sup>€</sup> | 0 (0%) |
| The Americas | 11 (30.6%)<sup>€</sup> | 2 (40%) |
| **Year of enrolment, studies (percentage)** |  |  |
| Pre-2015 | 6 (75%) | 5 (100%) |
| 2015-2019 | 2 (25%) | 0 (0%) |
| **Follow up duration in days, studies (percentage)** |  |  |
| 42 | 0 (0%) | 1 (20%) |
| >42 to <120 | 0 (0%) | 1 (20%) |
| 120 | 0 (0%) | 2 (40%) |
| >120 | 8 (100%) | 1 (20%) |
| **Age (weighted-average years)<sup>#,§</sup>** | 19.1 | 20.0<sup>§</sup> |
| **Male (weighted percentage)<sup>#</sup>** | 65.8% | 63.7% |

€ Included multinational studies. Number and percentage are derived from the number of study sites, not studies, within each region. # Weights approximated by the numbers of vivax patients treated with primaquine. \$ Study-specific average is the mean or median in vivax patients treated with primaquine. § Excluding one study for which the required summary statistics not available.

**Figure S3.**
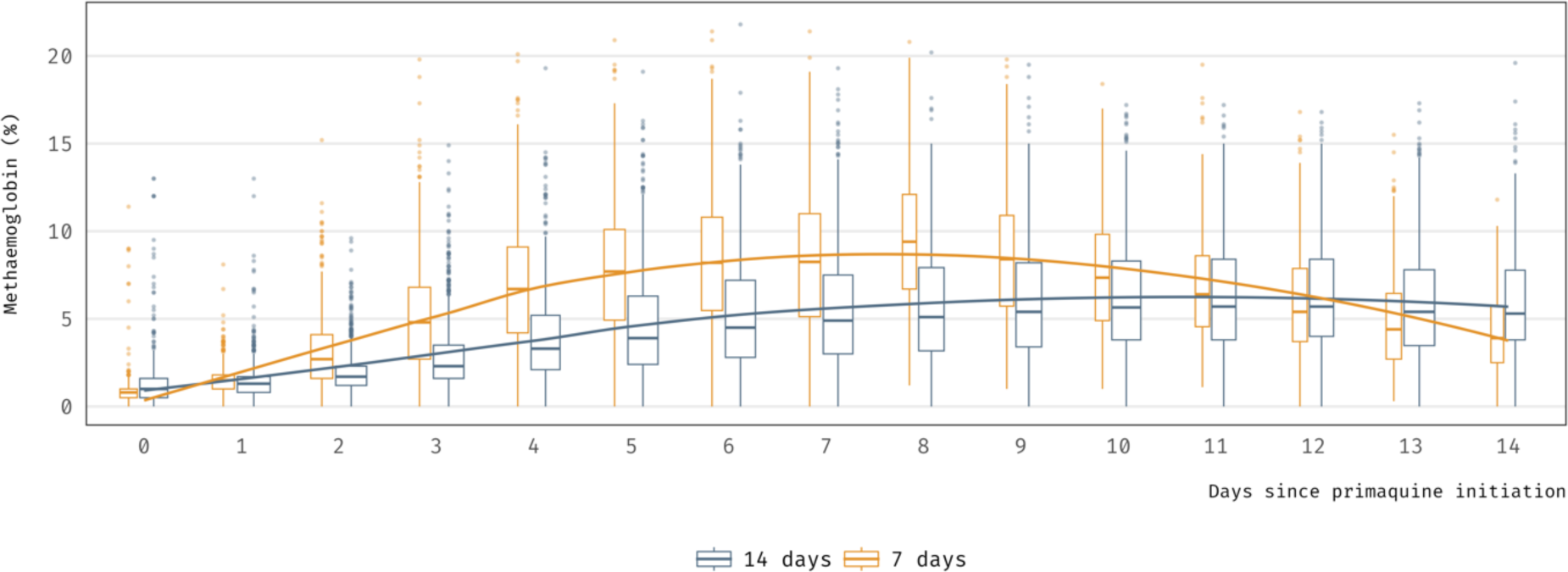
Dynamics of primaquine-induced increases in blood methaemoglobin over time. Box width is proportional to the square root of the number of patients.

**Figure S4.**
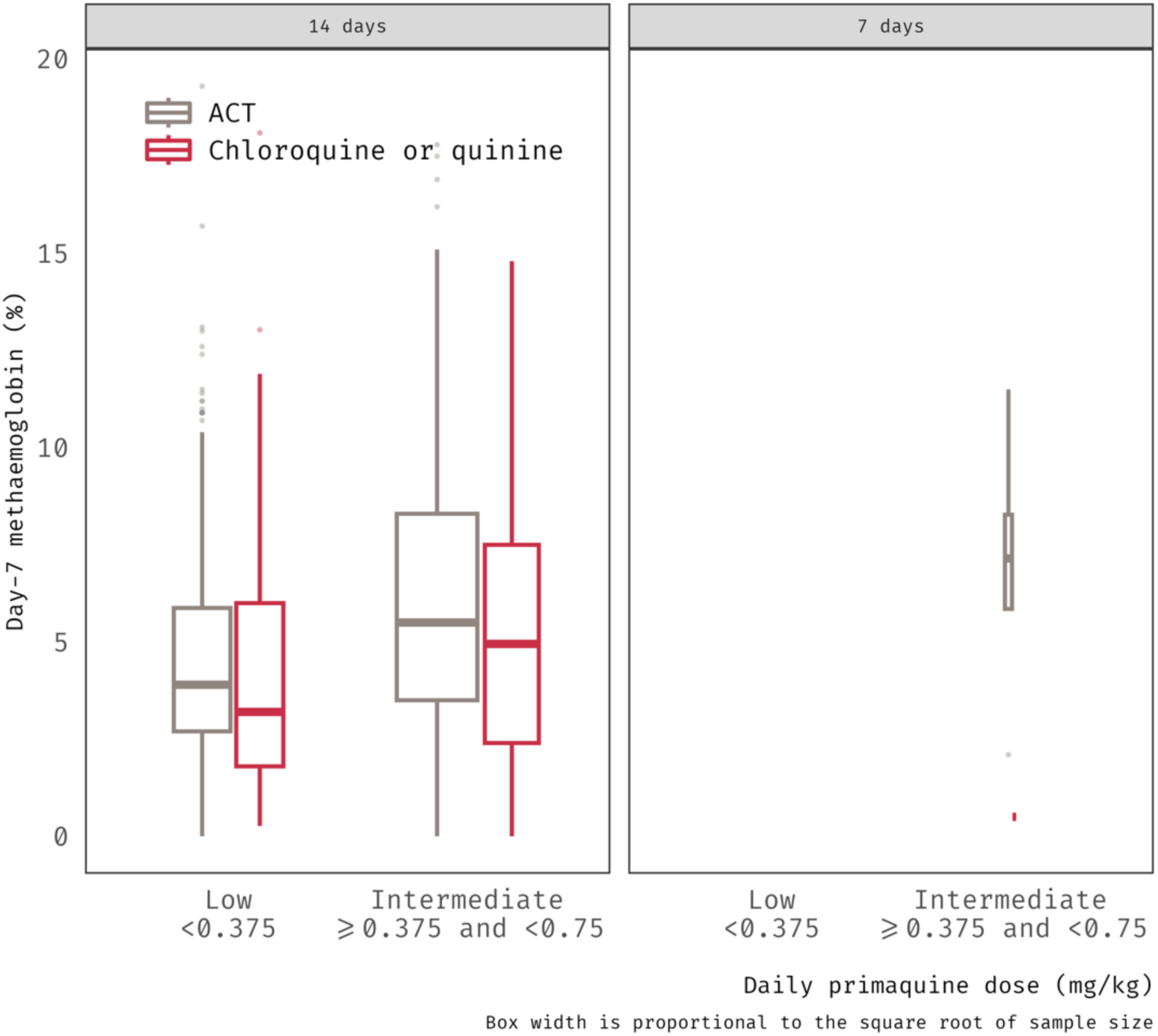
Day 7 methaemoglobin concentrations by primaquine regimen and dose group. Among patients treated with a low-to-intermediate daily primaquine dose, day 7 methaemoglobin was lower when primaquine was combined with chloroquine or quinine as a partner drug.

**Figure S5.**
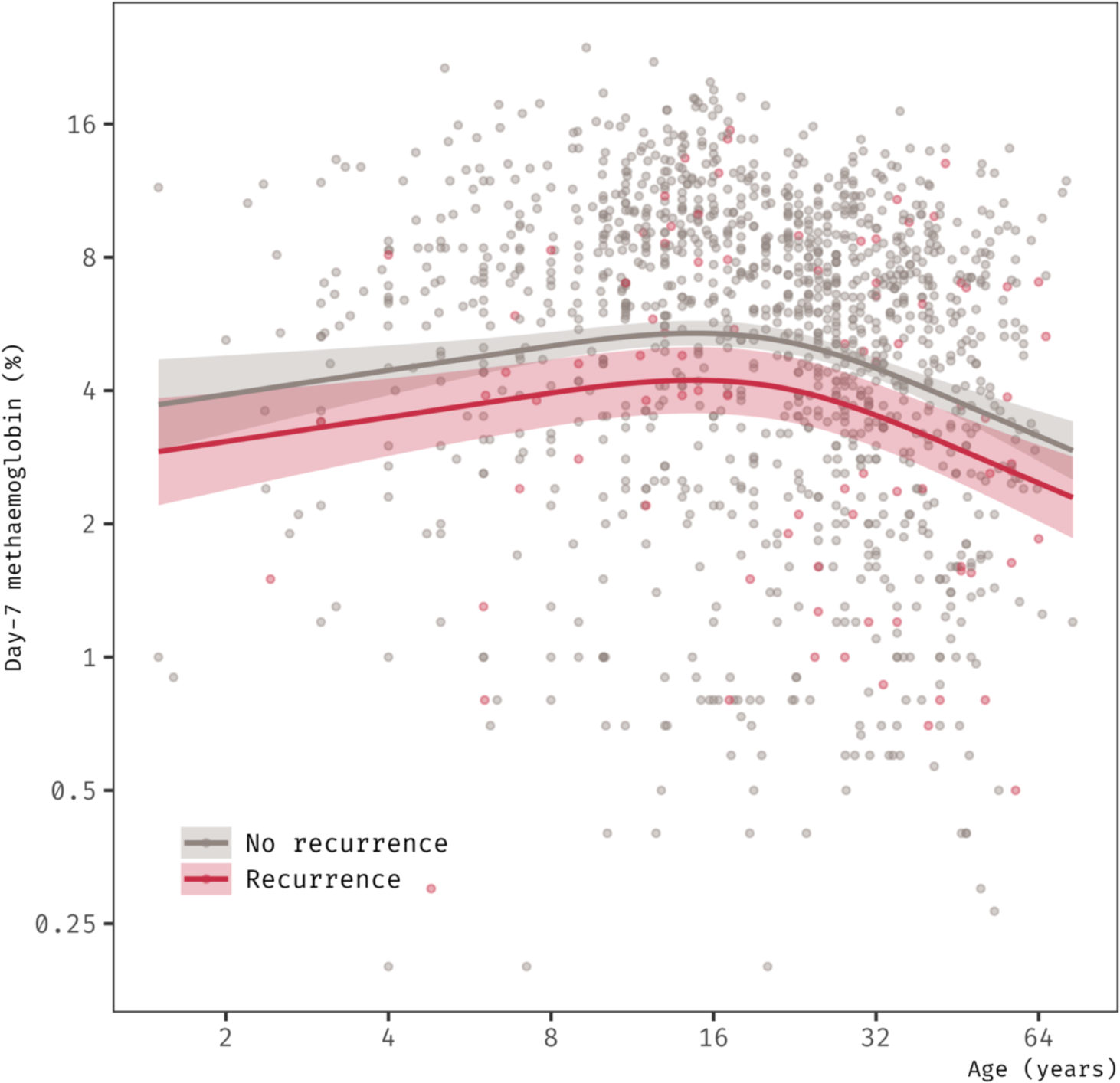
Inverse J-shaped association between patient age and day 7 methaemoglobin by recurrence status, after controlling for daily mg/kg primaquine dose. Horizontal and vertical axes are shown on the logarithmic scale.

**Figure S6.**
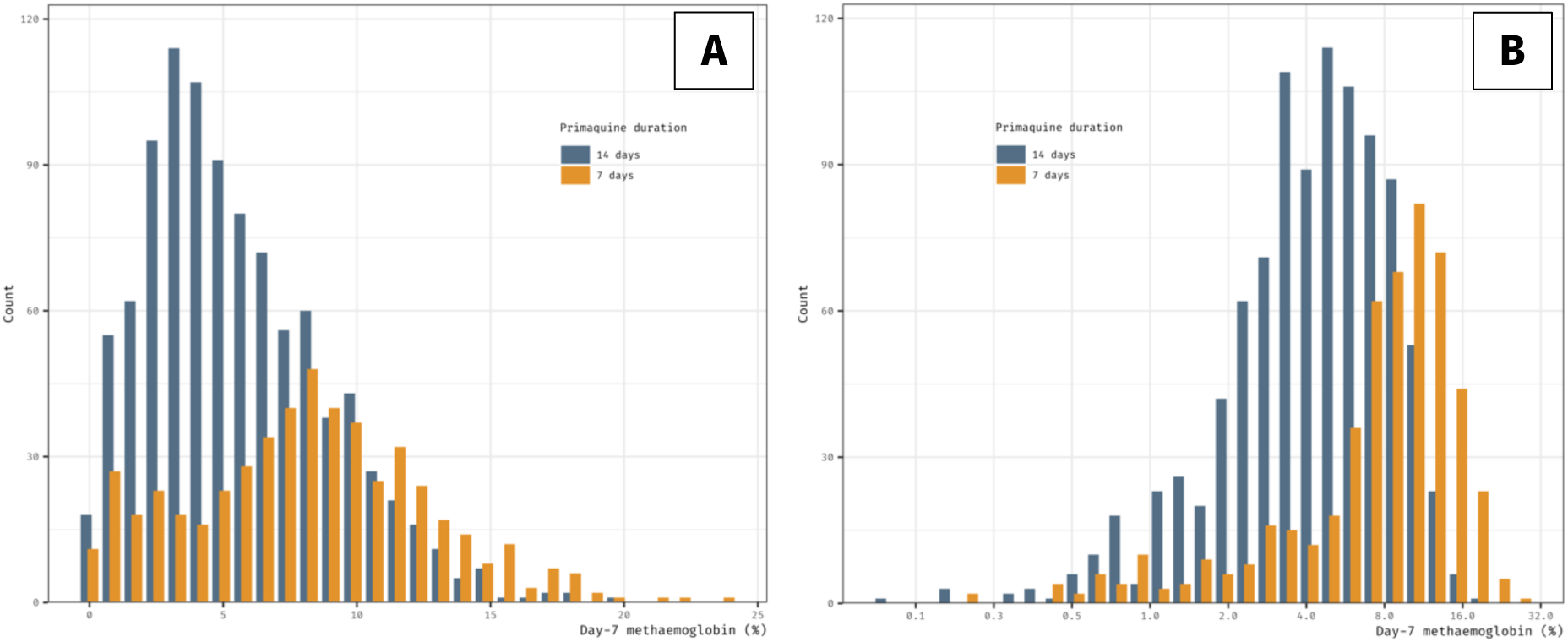
Distribution of day 7 methaemoglobin among the patients on (A) the original scale and (B) the logarithmic scale.

**Figure S7.**
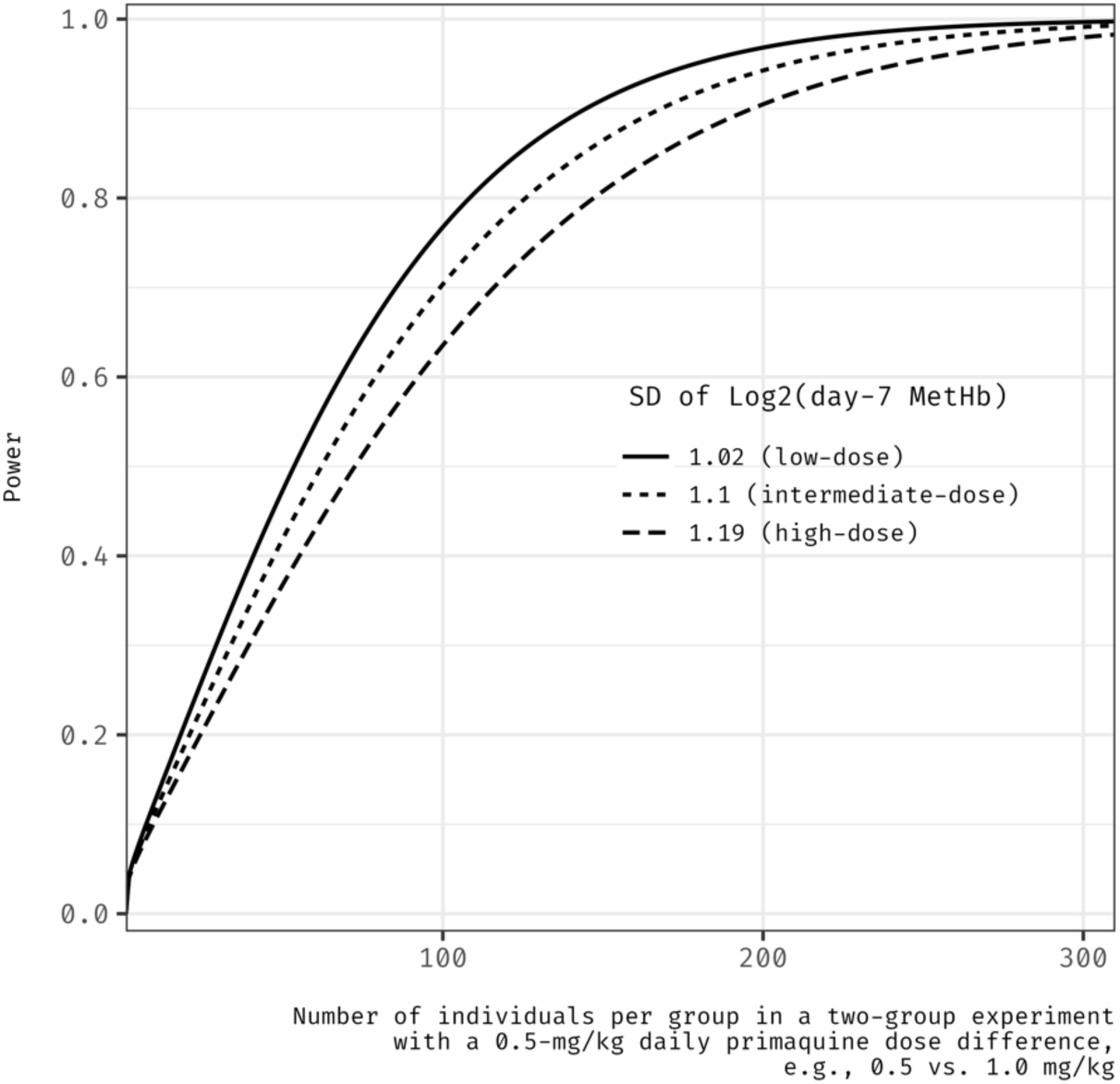
Example of sample size calculations for future studies. The assumed effect size is 0.39 (change on the log_2_ day 7 methaemoglobin) which is equivalent to a 0.5-mg/kg increase in daily primaquine dose. The standard deviation (SD) of the log_2_ day 7 methaemoglobin level were calculated for different categories of daily mg/kg primaquine dose based on pooled data. The false positive rate was set to 5%. The population distribution of the log_2_ of day 7 methaemoglobin level conditional on the daily dose was assumed to follow a normal distribution.

